# Assessing the impact of maternal blood pressure during pregnancy on perinatal health: A wide-angled Mendelian randomization study

**DOI:** 10.1101/2025.06.19.25329933

**Authors:** Fernanda Morales-Berstein, Ana Gonçalves-Soares, Qian Yang, Nancy McBride, Tom Bond, Marwa AL Arab, Alba Fernández-Sanlés, Maria C Magnus, Eleanor Sanderson, Emma Hart, Abigail Fraser, Katherine A Birchenall, Deborah A Lawlor, Gemma L Clayton, Maria-Carolina Borges

## Abstract

**Background:** Observational studies link high blood pressure in pregnancy to numerous adverse pregnancy and perinatal outcomes; however, findings may be affected by residual confounding or reverse causation. This study aimed to assess the causal effect of blood pressure during pregnancy on a range of pregnancy and perinatal outcomes.

**Methods:** We performed two-sample Mendelian randomization (MR) to assess the effect of systolic and diastolic blood pressure (SBP/DBP) during pregnancy on 16 primary and eight secondary adverse pregnancy and perinatal outcomes. We obtained genetic association data from large-scale meta-analyses of genome-wide association studies involving predominantly European ancestry individuals for SBP/DBP (N=1,028,980), and pregnancy and perinatal outcomes (N=77,285–934,566). We used inverse-variance weighted (IVW) MR for main analyses and MR-Egger, weighted median, weighted mode, multivariable MR, and IVW adjusted for fetal genetic effects for sensitivity analyses.

**Results:** A 10 mmHg higher genetically predicted maternal SBP increased the odds of gestational diabetes, induction of labour, low birth weight (LBW), small-for-gestational age (SGA), preterm birth (PTB), and neonatal intensive care unit (NICU) admission [OR ranging from 1.09 (95% CI: 1.04 to 1.14) for PTB to 1.33 (1.26 to 1.41) for LBW]; while decreasing the odds of high birth weight (HBW), large-for-gestational age (LGA), and post-term birth [OR ranging from 0.76 (0.69 to 0.83) for HBW to 0.94 (0.90 to 0.99) for post-term birth]. We did not find evidence that genetically predicted higher maternal SBP was related to miscarriage or stillbirth. Results for maternal DBP were similar to results for SBP. Overall, the main results were consistent across sensitivity analyses accounting for pleiotropic instruments and fetal genetic effects.

**Conclusions:** Higher maternal blood pressure reduces gestation duration and fetal growth, and increases the risks of induction of labour, gestational diabetes and NICU admission. This and other emerging evidence highlight the value of interventions to control maternal blood pressure during pregnancy to reduce the burden of adverse pregnancy outcomes.

## BACKGROUND

High blood pressure affects approximately one in 10 pregnant women, making it the most common medical problem encountered in pregnancy.^1^ Hypertensive disorders of pregnancy (HDP), which include chronic hypertension, gestational hypertension (GH) and preeclampsia, are among the leading causes of adverse pregnancy and perinatal outcomes worldwide,^2^ ^3^ especially in low- and middle-income countries.^4–6^ The burden of high blood pressure during pregnancy on maternal and offspring health is likely to increase with the rising prevalence of obesity and older maternal age at conception.^7–9^

There is an extensive body of literature from observational studies linking higher maternal blood pressure during pregnancy, as well as HDP, with a range of adverse pregnancy and perinatal outcomes, such as stillbirth, gestational diabetes (GDM), caesarean section, induction of labour, as well as having a baby who is small-for-gestational age (SGA), preterm (PTB), or requires admission to a neonatal intensive care unit (NICU).^10–13^ The evidence from these studies indicates that preventing or treating high blood pressure during pregnancy could help avert a wide range of adverse outcomes for mothers and babies. However, such studies are likely to be influenced by residual confounding due to unmeasured or inaccurately measured characteristics, such as socioeconomic position, maternal adiposity, and comorbidities. Furthermore, most observational studies to date have examined associations with only a limited number of pregnancy or perinatal outcomes. We argue that investigating a broader spectrum of these outcomes is crucial for informing more tailored blood pressure monitoring during pregnancy and for capturing the full scope of potential benefits associated with preventing hypertensive disorders in this period.

Evidence from randomised controlled trials (RCTs) on the effect of treating mild to moderate high blood pressure during pregnancy is uncertain for many adverse pregnancy and perinatal outcomes. A Cochrane systematic review on antihypertensive treatments including 5,909 women with mild to moderate hypertension during pregnancy (N=58 trials) was inconclusive on the treatment benefits for reducing adverse outcomes such as fetal or neonatal death (including miscarriage) (RR: 0.72, 95% CI 0.50 to 1.04), SGA (RR: 0.96, 95% CI: 0.78 to 1.18), and PTB (RR: 0.96, 95% CI: 0.83 to 1.12). ^14^ More recently, an open label RCT, enrolling 2,408 women with mild chronic hypertension, reported that being randomised to antihypertensive medications, as opposed to no treatment (unless severe hypertension developed), decreased the risk of a composite outcome, consisting of pre-eclampsia with severe features, medically indicated PTB, placental abruption and fetal/neonatal death.^14, 15^

Mendelian randomization (MR), a method that uses genetic variants as instrumental variables for modifiable risk factors, could help disentangle causation from confounding in an observational analysis.^16^ ^17^ MR can strengthen causal evidence to inform clinical decisions, especially when high-quality and well-powered RCTs are unavailable. Genetic variants are randomly allocated at conception and cannot be modified by the outcome of interest.

Hence, MR is less susceptible to reverse causation and confounding by factors such as socioeconomic position and related behavioural and health factors, than conventional observational methods.^18^ Previous MR studies have investigated the effects of higher genetically predicted maternal blood pressure on a limited set of outcomes, reporting potential effects on offspring birthweight^19^ ^20^ and gestational duration^21^ ^22^, although there were inconsistencies between studies, and some studies did not appropriately account for the correlation between maternal and fetal genotype.

We aimed to test the effect of higher blood pressure during pregnancy on a wider range of adverse pregnancy and perinatal outcomes. We did so by using MR to estimate the effect of genetically predicted higher systolic (SBP) and diastolic (DBP) blood pressure during pregnancy on 24 adverse maternal and offspring outcomes.

## METHODS

We examined the effect of maternal genetically predicted blood pressure on 16 primary and eight secondary pregnancy and perinatal outcomes using a two-sample MR framework (Figure 1). First, we selected genetic instruments for SBP (545 single-nucleotide polymorphisms (SNPs)) and DBP (513 SNPs) identified in a genome-wide association study (GWAS) meta-analysis by Keaton et al.^23^, including 1,028,980 European non-pregnant women and men (sample 1). Then, we extracted pregnancy and perinatal outcomes GWAS data for the same SNPs from up to 934,566 women from the MR-PREG collaboration (sample 2). Genetic association data from samples 1 and 2 were used to estimate the effect of maternal genetically predicted blood pressure (i.e., SBP and DBP) on the 24 pregnancy and perinatal outcomes. We conducted a series of sensitivity analyses to explore the plausibility of the core MR assumptions as outlined in Figure 1 and detailed under ‘Sensitivity analyses’. This study was reported following the recommendations of the STROBE-MR guidelines^25^ (Supplementary material).

**Figure 1.**
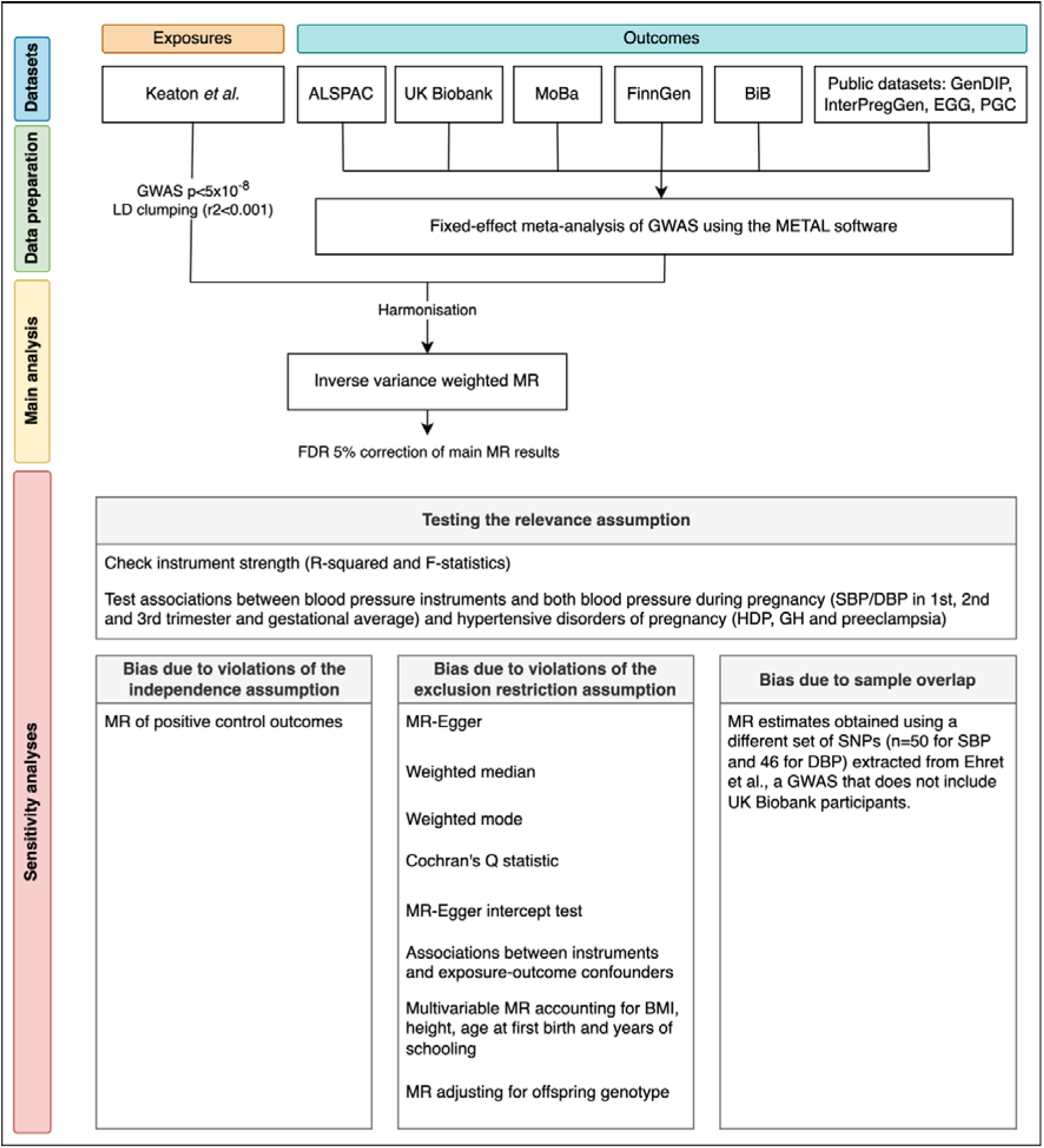
Flowchart for study design Abbreviations: ALSPAC, Avon Longitudinal Study of Parents and Children; BiB, Born in Bradford; BMI, body mass index; DBP, diastolic blood pressure; EGG, Early Growth Genetics consortium; FDR, false discovery rate; GenDIP, Genetics of Diabetes in Pregnancy consortium; GH, gestational hypertension; GWAS, genome-wide association study; HDP, hypertensive disorders of pregnancy; InterPregGen, International Pregnancy Genetics study; LD, linkage disequilibrium; MoBa, Norwegian Mother, Father and Child Cohort Study; MR, Mendelian randomization; PGC, Psychiatric Genetics Consortium; SBP, systolic blood pressure. Genetic association data for SBP and DBP were obtained from Keaton et al. (main analysis) and Ehret et al. (sensitivity analysis to assess bias due to sample overlap).

### Data sources

Details regarding the generation of genetic association data for blood pressure traits, perinatal outcomes, and data used in sensitivity analyses are provided in the Supplementary Methods.

We used genetic association data for SBP and DBP from the GWAS meta-analyses by Keaton et al.^23^ to maximise statistical power given this was the largest data source available at the time of the study (N=1,028,980 European women and men). For sensitivity analyses assessing bias due to sample overlap, we also selected SNPs from Ehret et al.^26^, a GWAS meta-analysis including 201,529 European women and men, who are not part of UK Biobank (UKB). In both GWAS, SBP and DBP were measured using standard protocols detailed elsewhere^23^ ^26^. For individuals treated with antihypertensive medication, measured values were adjusted by adding 15 and 10 mmHg to SBP and DBP, respectively, to account for antihypertensive treatment effects that could lead to the underestimation of the GWAS estimates^23^. Table 1 summarises key information on the GWAS used for SBP and DBP phenotypes.

**Table 1.** Description of publicly available GWAS used for SBP and DBP phenotypes.

| Study | Consortium (PMID) | Units | Sample size, n | European, % | UKB female participants, % | Sex | Covariate adjustments<sup>a</sup> | Accession codes |
| --- | --- | --- | --- | --- | --- | --- | --- | --- |
| Keaton 2024<sup>23</sup> | UKB + ICBP + MVP + BioVU (38689001) | mmHg | 1,028,980 | 100% | 24% | Male and female | Sex, age, age<sup>2</sup> and BMI | GCST90310294 (SBP)<br>GCST90310295 (DBP) |
| Ehret 2016<sup>26</sup> | ICBP (27618452) | mmHg | 201,529 | 100% | 0% | Male and female | Sex, age, age<sup>2</sup> and BMI | GCST006259 (SBP)<br>GCST006258 (DBP) |

<sup>a</sup>Additional adjustments: UKB was adjusted for genotyping chips. ICBP studies were optionally adjusted for population stratification using different methods (e.g., adjustment for principal components, adjustment for study site, multidimensional scaling). For more details, see Keaton et al.<sup>23</sup> and Ehret et al.<sup>26</sup> These data were downloaded from the NHGRI-EBI GWAS Catalog<sup>27</sup> data repository using the “MRInstruments” R package using the codes listed under the ‘Accession codes’ column.
Abbreviations: UKB, UK Biobank; ICBP, International Consortium for Blood Pressure; MVP, Million Veteran Program; BioVU, Vanderbilt University’s biorepository of DNA linked to de-identified medical records; SBP, systolic blood pressure; DBP, diastolic blood pressure; BMI, body mass index.

We were interested in 16 primary and eight secondary outcomes. Outcomes were classified as secondary if they were a subtype of or the continuous trait underlying a primary outcome. Our outcomes of interest included: 1) pregnancy loss outcomes including miscarriage (with sporadic and recurrent miscarriage as secondary outcomes) and stillbirth; 2) maternal morbidity outcomes including GDM and perinatal depression; 3) labour outcomes including induction of labour, pre-labour rupture of membranes and caesarean section (c-section, with emergency and elective c-section as secondary outcomes); 4) offspring birth outcomes including low and high birth weight (LBW/HBW) (with continuous birth weight in standard deviations as a secondary outcome), small- and large-for-gestational age (SGA/LGA), PTB and post-term birth (with continuous gestational age in weeks, and very PTB and spontaneous PTB as secondary outcomes), low Apgar score at 1 and 5 minutes, and NICU admission. Details on the outcome definitions can be found in Supplementary tables 1A (primary outcomes), 1B (secondary binary outcomes) and 1C (secondary continuous outcomes).

Outcome data were obtained from the MR-PREG collaboration,^24^ which included data from three birth cohorts — i.e., the Avon Longitudinal Study of Parents and Children (ALSPAC),^28^ ^29^ Born in Bradford (BiB)^30^ and the Norwegian Mother, Father and Child Cohort Study (MoBa),^31^ ^32^; two biobanks — i.e., UKB^33^ ^34^ and FinnGen;^35^ and four GWAS meta-analyses — i.e., the Early Growth Genetics (EGG) consortium,^36^ the International Pregnancy Genetics study (InterPregGen),^37^ the Genetics of Diabetes in Pregnancy (GenDIP) consortium^38^ and the Psychiatric Genomics Consortium (PGC).^39^

ALSPAC is a prospective birth cohort that recruited pregnant women from the former county of Avon, UK, between April 1991 and December 1992, enrolling 14,541 women and following their children over time.^28^ ^29^ The study collected extensive questionnaire data, biological samples, and anthropometric measurements, with genetic data available for mothers, partners, and children.

BiB is a prospective birth cohort that recruited 12,453 pregnant women in Bradford, UK, between 2007 and 2010, with most enrolled during their oral glucose tolerance test at 26– 28 weeks of gestation.^30^ The study collected biological samples, anthropometric data, and questionnaire responses, with linkage to primary and secondary care records and genetic data available for mothers and children.

MoBa is a nationwide Norwegian birth cohort that recruited over 95,200 mothers and 75,200 fathers between 1999 and 2008, with follow-up of 114,500 children.^31^ ^32^ The study collected questionnaire data, biological samples, and anthropometric measures, with linkage to the national Medical Birth Registry and genetic data available for families.

UKB is a large-scale adult cohort study that recruited 500,000 individuals aged 40–69 years across the UK between 2006 and 2010.^33^ ^34^ The study collected extensive baseline data, biological samples, and health measures, with ongoing follow-up via electronic health records and linkage to hospital and maternity admissions data.

FinnGen^35^ is a nationwide Finnish biobank network that integrates genetic data from 500,348 individuals with national electronic health registries, providing detailed information on prescriptions and disease diagnoses (ICD-9, ICD-10 codes). The study includes clinical endpoints, including adverse pregnancy and perinatal outcomes (APPOs), with case-control data available.

Where external, quality-controlled GWAS meta-analysis data from key genetic consortia were available, the MR-PREG collaboration^24^ harmonised these to supplement the sample size for the following outcomes: GDM from GenDIP (5,485 cases, 347,856 controls),^38^ preeclampsia from InterPregGen (4,630 cases, 373,345 controls),^37^ gestational duration traits from EGG (including PTB, post-term birth, and gestational age at delivery),^36^ and postnatal depression from PGC (17,339 cases, 53,426 controls).^39^

### Data analyses

1. Genetic instrument selection

We selected genetic instruments from the Keaton et al.^23^ GWAS to maximise statistical power. GWAS-significant SNPs (P<5×10^−8^) were selected as candidate instrumental variables for SBP and DBP. Among correlated SNPs, those with the smallest p-values were identified and retained via linkage disequilibrium (LD) clumping (r^2^<0.001, window size of 10,000 kb), using the 1000 Genomes European ancestry reference panel.

1. Genetic instrument relevance

We estimated instrument strength in the original SBP/DBP GWAS meta-analyses by calculating the F-statistic and R^2^ for the selected SNPs as follows: 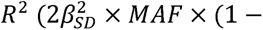 MAF)) ^40^ and 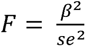 ^41^, where MAF is the minor allele frequency, N is the sample size and β is the effect estimate of the IV-blood pressure association with corresponding standard error (SE). We then computed the mean and range of the F-statistics, as well as the total R^2^ for each set of SBP/DBP instruments.

Due to the absence of large-scale GWAS meta-analyses of SBP and DBP measured during pregnancy, we selected SNPs from GWAS meta-analyses of non-pregnant individuals. This assumes that these SNPs are strong instruments for the exposure of interest, i.e., mean SBP and DBP during pregnancy. We tested this assumption in two steps.

First, we compared the SNP effects on SBP/DBP that were obtained using measurements of blood pressure taken among the general population (in mmHg)^23^ with those obtained using measurements taken during the first, second, and third trimesters, and across gestation (in mmHg using ALSPAC data). We calculated Pearson correlation coefficients to quantify these relationships. In addition, we ran linear regression models with SNP effect estimates from the general population as the independent variable and during pregnancy as the dependent variable. From these models, we extracted the regression intercept, slope, and R^2^. A slope of 1 implies that the SNPs have the same average effect in both ALSPAC and the general population; a slope<1 implies smaller effects in ALSPAC while a slope>1 implies larger effects in ALSPAC.

Second, we checked whether there was a positive correlation between the SNP effects on SBP/DBP in the general population (from Keaton et al.^23^) and the SNP effects on the risk of HDP (cases/N=32,549/541,768), GH (cases/N=20,777/527,932), and preeclampsia (cases/N=19,566/678,001) among the participants included in the MR-PREG meta-analyses.

1. Data harmonisation

Genetic association data for SBP and DBP were harmonised with those for perinatal outcomes to ensure effect alleles matched across datasets. Positive strands of palindromes were aligned using minor allele frequencies (MAF). Palindromic SNPs with MAF ≥ 0.42 or strand mismatch were excluded.

1. MR analyses

### Main analyses

The main MR analyses were performed using the inverse-variance weighted (IVW) method^42^, which has the greatest statistical power. MR estimates were scaled to reflect a 10-mmHg increase in genetically predicted SBP and DBP (this was also done in the sensitivity MR analyses described below).

A 5% Benjamini-Hochberg False Discovery Rate (FDR) correction^43^ was applied to all main IVW results to account for multiple testing.

### Sensitivity analyses

We conducted a series of sensitivity analyses to assess the robustness of our main MR estimates. These were mainly focused on assessing the plausibility of the core assumptions required for MR to reliably test the presence of a causal effect: genetic instruments must 1) be strongly statistically associated with blood pressure during pregnancy, 2) share no common cause with the perinatal outcomes of interest, and 3) only be associated with the perinatal outcomes via their effect on maternal blood pressure (e.g., no unbalanced horizontal pleiotropy)^44^. These are generally referred to as the relevance, independence, and exclusion restriction assumptions, respectively. We tested the relevance assumption as described above under ‘Genetic instrument relevance’. For the independence and exclusion restriction assumptions, we assessed their plausibility through the sensitivity analyses described below.

a. Bias due to violations of the independence assumption

The independence assumption of no confounding between the genetic instrument and outcomes could be violated by population stratification, assortative mating, or dynastic effects.^45^ For biological traits such as blood pressure, assortative mating and dynastic effects are less likely to be key sources of bias.^46^ To mitigate bias due to population stratification, all GWASes used in our analyses were adjusted for principal components of ancestry and/or used mixed models.

We also conducted MR analyses assessing the effects of genetically predicted SBP/DBP on positive (i.e., stroke and coronary artery disease) control outcomes. Genetic association data for these outcomes were accessed through the IEU OpenGWAS database API (IDs: ebi-a-GCST005838 and ebi-a-GCST003116, respectively)^47^ ^48^.

Bias due to violations of the exclusion restriction assumption

We reran the analyses using MR methods that are understood to be more robust in the presence of horizontal pleiotropy: MR-Egger^49^, weighted median,^50^ and weighted mode^51^ methods. All of these rely on different assumptions about the nature of any pleiotropic effects. MR-Egger assumes instrument-outcome pleiotropic effects are independent of instrument-risk factor associations (Instrument Strength Independent of Direct Effect — InSIDE — assumption). The slope of the MR-Egger regression represents the estimate of the true causal effect, if the InSIDE^49^assumption holds. The weighted median estimator ^50^ assumes that at least 50% of the weight in the analyses comes from valid instruments. The weighted mode estimator assumes that the most frequent effect equals the true causal effect (ZEro Modal Pleiotropy Assumption — ZEMPA).^51^ Consistent results across methods are supportive of MR estimates being unbiased.

Cochran’s Q statistic was used to quantify heterogeneity across estimates obtained from different SNPs^52^. A large Q statistic is indicative of high levels of heterogeneity in the individual SNP estimates, possibly but not necessarily due to horizontal pleiotropy.

Importantly, this test is not specific to violations of the exclusion restriction criteria but can indicate that one or more genetic variants violate the MR assumptions. Additionally, MR-Egger intercept tests were performed to assess statistical evidence for unbalanced horizontal pleiotropy. A non-zero intercept (p<0.05) can be interpreted as evidence of unbalanced pleiotropic bias.

To investigate potential specific pleiotropic pathways (via established risk factors for adverse perinatal outcomes), we also conducted MR analyses assessing the effects of genetically predicted SBP and DBP on height (id: ieu-b-4814), body mass index (BMI; id: ieu-b-4816), age at first birth (id: ieu-b-4820), age at menarche (id: ieu-b-4822), cigarettes smoked per day (id: ieu-b-4826), number of children ever born (id: ieu-b-4828), alcohol consumption (id: ieu-b-4834), years of schooling (id: ieu-b-4836), and physical activity (id: ieu-b-4860) in the UKB.^53^ Details of how these risk factors were assessed are provided in the Supplementary Methods. Any evidence of an effect would suggest potential violations of the exclusion restriction assumption. We used multivariable MR^54^ to account for any potentially pleiotropic risk factors (i.e., that were predicted by our genetic instruments for SBP/DBP) by including them as additional exposures in each genetically instrumented maternal blood pressure-perinatal outcome model. The multivariable MR exposure and outcome datasets were all harmonised to the same effect allele, and we calculated the conditional F-statistics to test whether the main exposure could be strongly predicted in the multivariable MR.

Furthermore, MR analyses adjusted for offspring genotype were conducted to investigate whether MR estimates were biased by fetal effects of maternally inherited alleles (which would lead to the violation of the exclusion restriction assumption). We accounted for a potential effect from fetal genetic variants using a weighted linear model (WLM).^5556^ First, we used a WLM to estimate conditional genetic effects — i.e., maternal genetic effects on outcomes adjusted by offspring genotype. Second, we used IVW, as in the main analyses, with conditional estimates for maternal genetic effects on each outcome. We conducted additional analyses in which we simultaneously adjusted for offspring and paternal genetic genotypes, whenever available, due to the potential collider bias related to only conditioning on fetal genotype.^57^

Bias due to sample overlap

Bias due to sample overlap between exposure and outcome datasets was assessed by comparing IVW MR estimates from partially overlapping datasets (Keaton et al.^23^) with those from non-overlapping datasets (Ehret et al.^26^).

1. Software

All analyses were conducted using R version 4.4.0. Two-sample MR analyses were performed using the “TwoSampleMR” package version 0.6.6. Multivariable MR analyses were conducted using the “MVMR” package version 0.4 available at https://github.com/WSpiller/MVMR. The WLM models were implemented in the DONUTS R package version 1.0.0.^56^ Plots were created using “ggplot2” package version 3.5.1 and the “ggforestplot” package version 0.1.0.

## RESULTS

### Descriptive results

The total number of participants (and cases) for each outcome included in the present MR study is shown in Table 2. The number of participants ranged from 77,285 (NICU admission) to 934,566 (GDM), while the number of cases ranged from 876 (low Apgar score at 5 minutes) to 89,086 (miscarriage).

**Table 2.** Total number of participants included in the study by outcome of interest.

| Outcome | Type | Cases (N) | Cases (%) | N |
| --- | --- | --- | --- | --- |
| ***Pregnancy loss outcomes*** |  |  |  |  |
| Miscarriage | primary | 89,086 | 18% | 397,131 |
| Stillbirth | primary | 6,331 | 3% | 201,339 |
| Sporadic miscarriage | secondary | 55,888 | 23% | 185,271 |
| Recurrent miscarriage | secondary | 6,233 | 2% | 321,756 |
| ***Maternal morbidity outcomes*** |  |  |  |  |
| Gestational diabetes | primary | 26,346 | 3% | 934,566 |
| Perinatal depression | primary | 22,785 | 15% | 147,601 |
| ***Labour outcomes*** |  |  |  |  |
| Induction of labour | primary | 14,495 | 15% | 95,239 |
| Rupture of membranes | primary | 21,839 | 7% | 310,621 |
| Caesarean section | primary | 33,909 | 14% | 233,969 |
| Emergency C-section | secondary | 9,165 | 10% | 90,438 |
| Elective C-section | secondary | 6,004 | 7% | 87,283 |
| **Offspring birth outcomes** |  |  |  |  |
| Low birth weight | primary | 19,617 | 7% | 275,214 |
| High birth weight | primary | 6,679 | 3% | 266,835 |
| Small for gestational age | primary | 6,882 | 8% | 89,033 |
| Large for gestational age | primary | 10,468 | 11% | 96,305 |
| Preterm birth | primary | 20786 | 5% | 412,435 |
| Post-term birth | primary | 27,841 | 7% | 425,673 |
| Low Apgar score at 1 minute | primary | 4,950 | 6% | 86,668 |
| Low Apgar score at 5 minutes | primary | 876 | 1% | 78,739 |
| NICU admission | primary | 6,996 | 9% | 77,285 |
| Spontaneous preterm birth | secondary | 6629 | 5% | 147,055 |
| Very preterm birth | secondary | 1,214 | 1% | 85,274 |
| Birth weight | secondary | NA | NA | 289,846 |
| Gestational age | secondary | NA | NA | 226,810 |

Abbreviations: NICU, neonatal intensive care unit; NA, not applicable. N, maximum number of participants (it may be lower for some genetic variants).

### Genetic instruments relevance

We selected 545 and 513 independent GWAS-significant SNPs (P<5×10^-8^) as instrumental variables for SBP and DBP, respectively. Altogether, they each explained 3.5% and 4.4% of the variance in SBP and DBP. Mean F-statistics across SNPs corresponded to 80 and 82 for the SBP and DBP instruments, respectively. Genetic instruments used in the analyses are detailed in Supplementary Tables 2-5.

The selected SNPs were associated with SBP/DBP during pregnancy and with the risk of HDP, GH, and preeclampsia (correlations are shown in Supplementary Figures 1a to 1e and the regression intercept, slope, and R^2^ are shown in Supplementary Figures 2a to 2e). For illustration, there was a positive correlation between the effect of the blood pressure increasing alleles on SBP in the general population and their effect on SBP among study participants in their first (Pearson’s coefficient (r) = 0.34, p = 4.4×10^-16^), second (r = 0.39, p = 2.2×10^-16^), and third trimesters of pregnancy (r = 0.39, p = 2.2×10^-16^), as well as across their full gestation (r = 0.42, p = 2.2×10^-16^). Corresponding correlations for DBP ranged from 0.19 (p = 1.3×10^-5^) to 0.49 (p = 2.2×10^-16^). Similarly, the regression models to examine the association of SNPs with SBP and DBP throughout the gestational period had, respectively, intercepts of −0.01 and 0.01, and slopes of 0.34 (p=9.7×10^-25^) and 0.45 (p=1.9×10^-30^). In addition, there was a positive correlation between the effect of blood pressure increasing alleles on SBP in the general population and their effect on the risk of HDP (r = 0.76, p < 2.2×10^-16^), GH (r = 0.75, p < 2.2×10^-16^), and preeclampsia (r = 0.64, p < 2.2×10^-16^). Corresponding correlations for DBP ranged from 0.64 (p < 2.2×10^-16^) to 0.73 (p < 2.2×10^-16^).

### Main Mendelian randomization results for primary outcomes

Higher genetically instrumented maternal SBP was related to higher odds of GDM (OR=1.11, 95%CI: 1.05 to 1.17, per 10 mmHg), induction of labour (OR=1.11, 95%CI 1.04 to 1.17), LBW (OR=1.33, 95%CI 1.26 to 1.41), SGA (OR=1.16, 95%CI 1.07 to 1.27), PTB (OR=1.09, 95%CI 1.04 to 1.14), and NICU admission (OR=1.11, 95%CI 1.02 to 1.20), and lower odds of HBW (OR=0.76, 95%CI 0.69 to 0.83), LGA (OR=0.87, 95%CI 0.80 to 0.94), and post-term birth (OR=0.94, 95%CI 0.90 to 0.99). All these results passed our criteria for multiple testing correction. We did not find evidence that higher maternal SBP was related to miscarriage (OR=1.00, 95%CI 0.98 to 1.02) or stillbirth (OR=1.00, 95%CI 0.93 to 1.08). For other outcomes, the results were inconclusive due to the modest magnitude and relatively high imprecision of the effect estimates – i.e., perinatal depression (OR=1.03, 95%CI 0.98 to 1.09), and low Apgar at 1 (OR=1.04, 95%CI 0.94 to 1.14) and 5 minutes (OR=1.06, 95%CI 0.87 to 1.30) (Figure 2A). Results for maternal DBP were broadly similar to results for SBP, although they were estimated with higher imprecision (Figure 2B). MR estimates can also be found in Supplementary Table 6. Study-specific estimates are displayed in Supplementary Figure 3.

**Figure 2.**
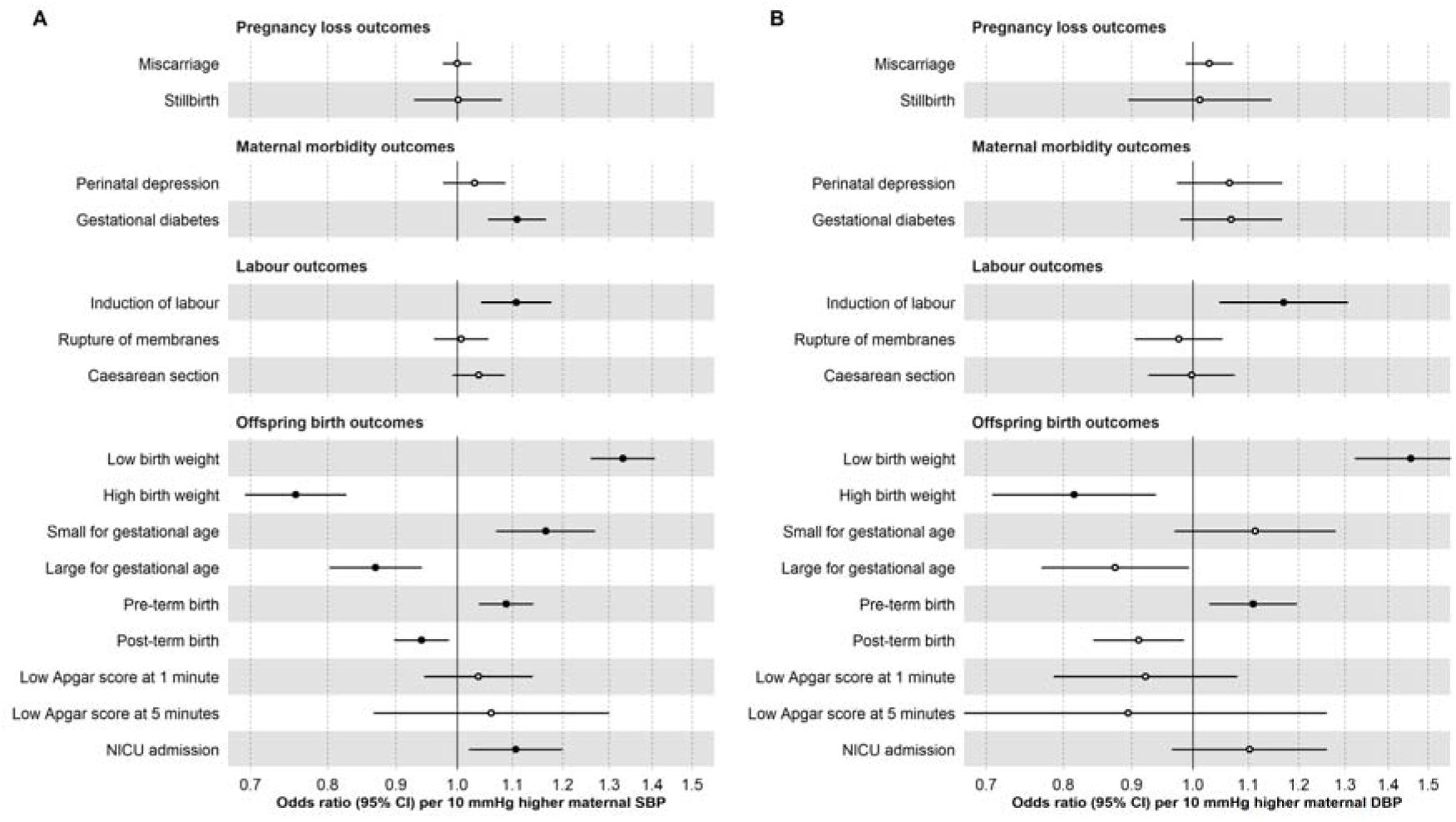
Inverse-variance weighted Mendelian randomization estimates for genetically predicted effects of maternal blood pressure on the primary outcomes. Odds ratios and 95% confidence intervals are presented per 10 mmHg increase in (A) systolic and (B) diastolic blood pressure. Systolic and diastolic blood pressure were instrumented by 545 and 513 genetic variants obtained from Keaton et al., respectively. Estimates with a false discovery rate corrected p-value <0.05 are indicated by filled black circles. Abbreviations: SBP, systolic blood pressure; DBP, diastolic blood pressure; NICU, neonatal intensive care unit.

### Main Mendelian randomization results for secondary outcomes

Consistent with findings from the primary outcomes, higher maternal SBP was related to secondary outcomes in a similar pattern: increased odds of emergency and elective C-section, spontaneous and very PTB, but not pregnancy loss outcomes (sporadic and recurrent miscarriage) (Supplementary Figure 4A). Additionally, higher SBP was linked to lower birthweight and shorter gestational period (Supplementary Figure 5A). Higher maternal DBP was associated with increased odds of adverse labour outcomes (elective C-section only), offspring outcomes (spontaneous and very PTB, lower birthweight, and shorter gestational age) (Supplementary Figures 4B and 5B). In contrast to the main results regarding any/sporadic miscarriage, higher DBP was related to higher odds of recurrent miscarriage (Supplementary Figure 4B). Combined results can also be found in Supplementary Table 6. Study-specific estimates are displayed in Supplementary Figures 6-7.

### Sensitivity analyses

There was some evidence of heterogeneity between SNP estimates for 12/16 primary outcomes in SBP analyses and 11/16 primary outcomes in DBP analyses (Cochran’s Q = 396-509, p<0.001) (Supplementary Table 7).

We found evidence that a higher genetically predicted SBP/DBP increased the risk of positive control outcomes (i.e., stroke and coronary artery disease) (Supplementary Figure 8).

The MR-Egger intercept test suggested that there was detectable evidence of unbalanced horizontal pleiotropy for SBP with the odds of induction of labour (intercept: −0.004, p = 0.02) and low Apgar score at 5 minutes (intercept: −0.014, p = 0.03). For DBP, there was detectable evidence with GDM (intercept: 0.005, p = 0.01), HBW (intercept: 0.007, p = 0.02), LGA (intercept: 0.007, p = 0.01), and low Apgar score at 5 minutes (intercept: −0.014, p = 0.03) (Supplementary Table 8).

As observed in Figure 3 (primary outcomes) and Supplementary Figures 9-10 (secondary outcomes), the direction of the MR estimates from the main analyses using IVW was consistent with the direction of the MR estimates in sensitivity analyses exploring potential bias due to unbalanced pleiotropy using MR-Egger, weighted median, and weighted mode methods. In some instances, the MR-Egger and/or the weighted mode estimates were attenuated and had larger confidence intervals (e.g., GDM, LGA, PTB, post-term birth, and NICU admission).

**Figure 3.**
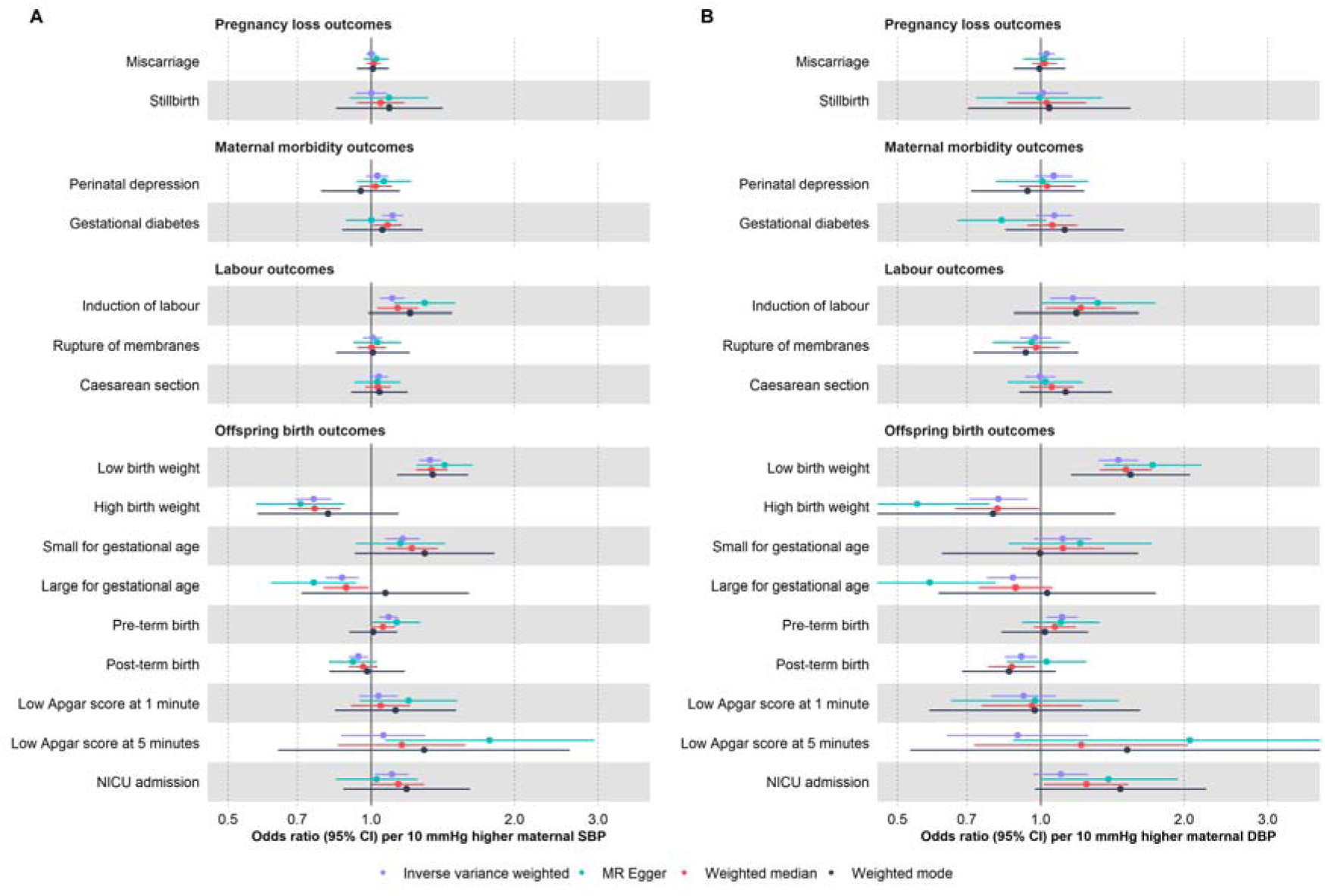
Mendelian randomization estimates for genetically predicted effects of maternal blood pressure on the primary outcomes across different methods. Odds ratios and 95% confidence intervals are presented per 10 mmHg increase in (A) systolic and (B) diastolic blood pressure. Systolic and diastolic blood pressure were instrumented by 545 and 513 genetic variants obtained from Keaton et al., respectively. Abbreviations: SBP, systolic blood pressure; DBP, diastolic blood pressure; NICU, neonatal intensive care unit.

We further investigated potential bias due to pleiotropic pathways. First, we checked whether higher blood pressure, as instrumented by our selected SNPs, affected well-established determinants of adverse pregnancy/perinatal outcomes. We found that higher genetically predicted blood pressure, particularly SBP, was associated with lower height, BMI, younger age at first birth, and fewer years of schooling (Supplementary Figure 11). The association of blood pressure increasing alleles with lower BMI is likely explained by the use of BMI-adjusted GWAS summary statistics for SBP/DBP, which has been shown to potentially lead to collider stratification bias in MR.^58^ ^59^ Second, we used multivariable MR to account for putative colliders (BMI) and pleiotropic factors (height, younger age at first birth, and fewer years of schooling). We found that the direction of the effect estimates remained unchanged and that their magnitude was only marginally affected (Figure 4, Supplementary Figure 12-13). The conditional F-statistics for SBP and DBP in the multivariable MR analyses ranged from 15 to 34, suggesting that it is unlikely that the results were affected by substantial weak instrument bias.

**Figure 4.**
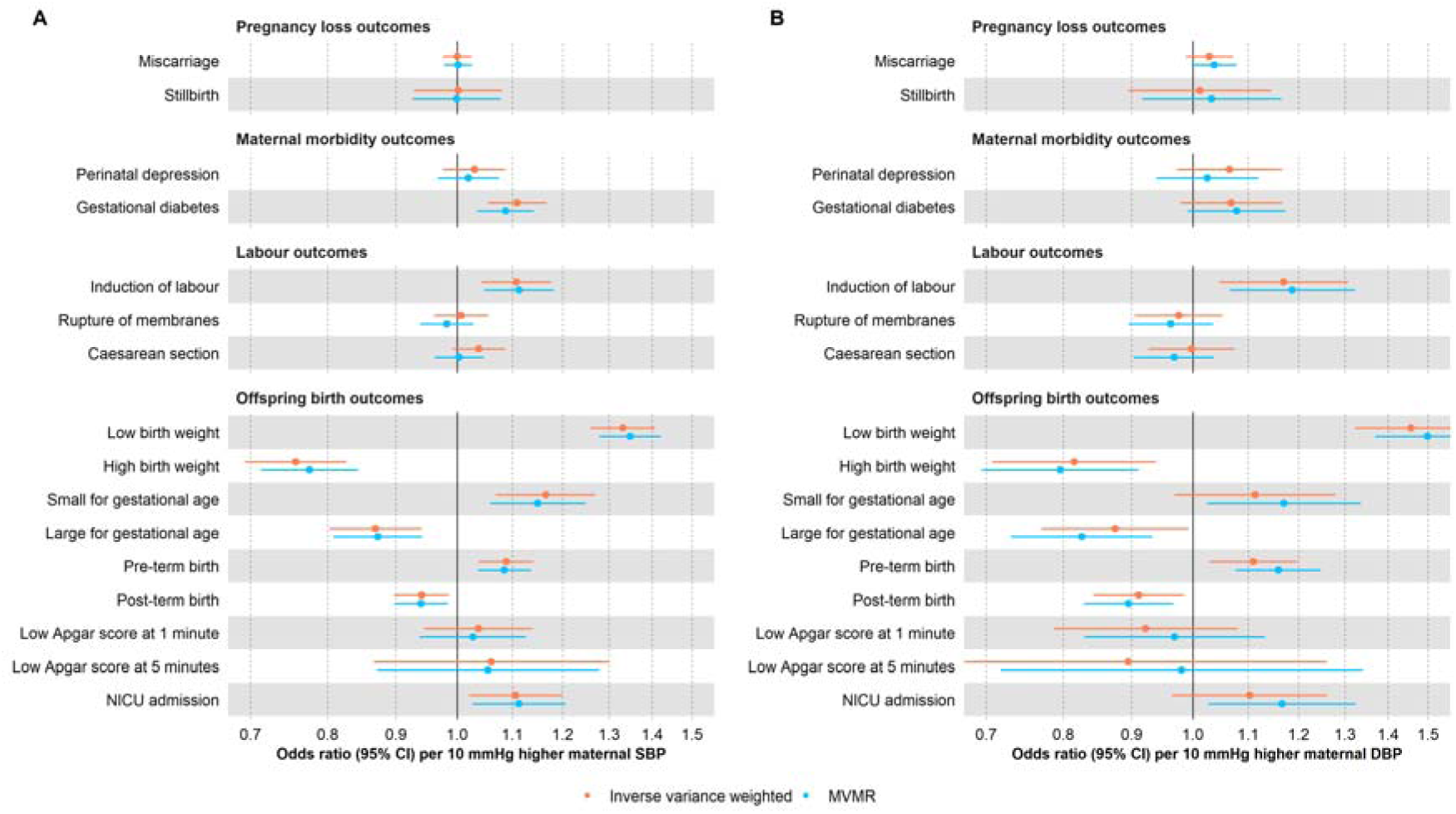
Univariable (“Inverse variance weighted”) and multivariable (“MVMR”) Mendelian randomization estimates for genetically predicted effects of maternal blood pressure on the primary outcomes. Multivariable models have been adjusted for body mass index, height, years of education, and age at first birth. Odds ratios and 95% confidence intervals are presented per 10 mmHg increase in (A) systolic and (B) diastolic blood pressure. Systolic and diastolic blood pressure were instrumented by 545 and 513 genetic variants obtained from Keaton et al., respectively. Abbreviations: SBP, systolic blood pressure; DBP, diastolic blood pressure; NICU, neonatal intensive care unit; MVMR: multivariable Mendelian randomization.

Additionally, in intergenerational analyses such as the present study, there could be bias by fetal genetic effects – i.e., where fetal genotype, inherited from the mother, affects the outcomes of interest. To test whether this was likely to be a source of bias in our main findings, we conducted MR analyses that accounted for offspring genotype. The direction of the effect estimates accounting for offspring genotype was generally consistent with the direction of the effect estimates in the main analysis, both for primary (Figure 5) and secondary outcomes (Supplementary Figures 14-15). For outcomes related to fetal growth (e.g., SGA and LGA), adjusting for fetal genetic effects led to the partial attenuation of effect estimates (Figure 5 and Supplementary Figure 15).

**Figure 5.**
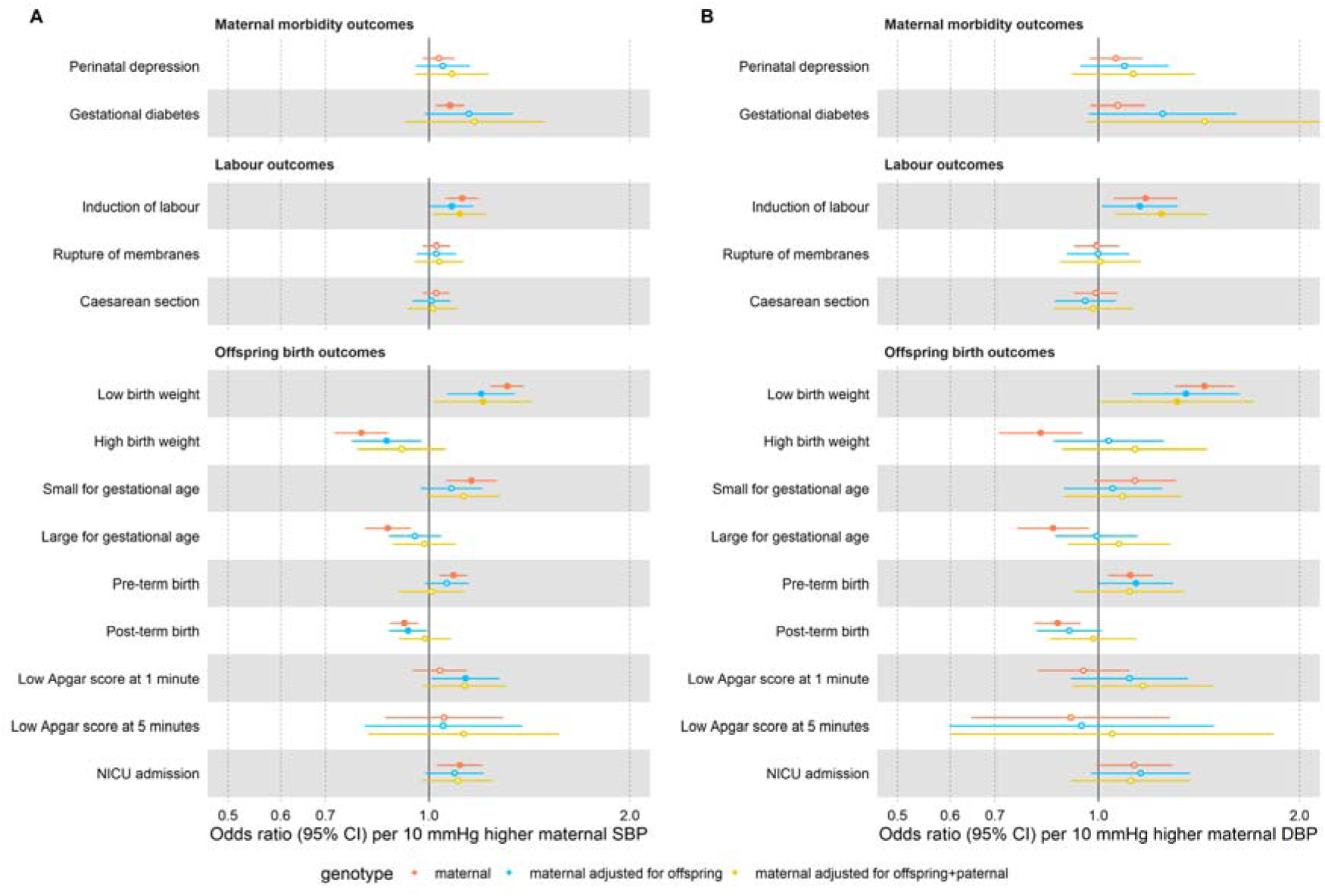
Mendelian randomization estimates for genetically predicted effects of maternal blood pressure on the primary outcomes, adjusting for offspring genotype. Odds ratios and 95% confidence intervals are presented per 10 mmHg increase in (A) systolic and (B) diastolic blood pressure. Estimates are shown for maternal effects unadjusted (red) and adjusted by fetal genetic effects (blue), and adjusted for both offspring and genetic effects (yellow). Systolic and diastolic blood pressure were instrumented by 545 and 513 genetic variants obtained from Keaton et al., respectively. Abbreviations: SBP, systolic blood pressure; DBP, diastolic blood pressure; NICU, neonatal intensive care unit.

Finally, we did not observe any evidence of bias due to sample overlap; the results from the non-overlapping samples (Ehret et al.^26^ sensitivity analyses) are consistent with those from the overlapping samples (Keaton et al.^23^ main analyses) (Supplementary Figures 16-18).

## DISCUSSION

Our findings align with a substantial body of observational research suggesting that elevated blood pressure during pregnancy impairs fetal growth and shortens gestational duration.^10^ ^11^ ^13^ The estimated effect of higher blood pressure on the risk of PTB may be partly explained by hypertension being a common reason for induction and medically-indicated PTB. However, it is important to note that we also observed a potential effect of higher maternal blood pressure on the risk of spontaneous PTB. Additionally, our results add to existing evidence that elevated blood pressure during pregnancy may increase the risk of GDM and NICU admission. Nevertheless, our results contrast with those from previous observational studies, which found that higher blood pressure during pregnancy increases the risk of miscarriage and stillbirth.^10^ ^13^

During pregnancy, blood pressure initially declines, before progressively increasing towards term. In a study including 4,607 women with low-risk pregnancies, median SBP was lowest at 12 weeks (111.5 mmHg, 95% CI: 111.3 to 111.8), rising to maximum at 40 weeks of gestation (119.6, 118.9 to 120.3 mmHg). Median DBP was lowest at 19 weeks of gestation (68.5, 68.3 to 68.7) and peaked at 40 weeks of gestation (76.3, 75.9 to 76.8 mmHg).^60^ Normal ranges for blood pressure not only vary with gestational age but also by maternal characteristics (nulliparous vs multiparous, pre-pregnancy BMI, and smoking status).^61^ This early decline in blood pressure reflects a normal physiological adaptation to pregnancy, driven by a reduction in systemic vascular resistance. This reduction is primarily due to the development of low resistance in uteroplacental circulation and hormonal influences, including the increased production of vasodilatory hormones such as relaxin.^62^ Based on the expected decline in blood pressure during the first 20 weeks of gestation, evidence of hypertension before this period is classified as chronic hypertension, and after this period as new-onset hypertension (i.e., gestational hypertension or preeclampsia).^63^ Despite the dynamic nature of blood pressure across gestation, we showed that our selected genetic instruments were associated with similar differences in SBP and DBP across trimesters.

Existing evidence indicates that greater increases in maternal blood pressure from early (18 weeks) to late (36 weeks) pregnancy are related to reduced fetal growth and shorter gestation, even among women who remained normotensive throughout pregnancy and did not meet the criteria for HDP.^64^ These associations are plausibly mediated by impaired oxygen and nutrient delivery to the fetus,^65^ as elevated blood pressure can disrupt key physiological processes at various stages of pregnancy, e.g., early placentation and subsequent placental perfusion.^66^ Interestingly, a previous study using haplotype-based genetic scores suggested that maternal blood pressure effects on offspring birthweight were biased by offspring genetic effects.^22^ In our study, we observed only partial attenuations when accounting for offspring genetic effects, indicating that maternal blood pressure has a causal effect on offspring birthweight, as corroborated by other studies.^20^ Our findings also suggest that higher genetically predicted maternal blood pressure is associated with an increased risk of labour induction, as expected given that HDP are a known indication for induction.^67^ Additionally, we observed a higher risk of NICU admission, with prior research identifying PTB, respiratory disease, and hypoglycaemia as the most common primary reasons for admission linked to maternal hypertension.^68^

Unlike conventional observational studies,^10^ ^13^ we were unable to confirm an effect of maternal blood pressure on miscarriage and stillbirth, despite observing an impact on fetal growth restriction and preeclampsia. Two plausible explanations for these findings are that previous studies may have overestimated the effect due to residual confounding (e.g., by factors that may independently affect maternal blood pressure and stillbirth, such as smoking or placental factors), or that effects are only present at more extreme values of blood pressure which may not be adequately captured by our genetic instruments since these generally explain only a small proportion of blood pressure variation. Our observation that higher blood pressure during pregnancy was related to higher risk of GDM is in line with conventional observational and MR studies’ finding of a positive link between blood pressure and the risk of type 2 diabetes. However, it is uncertain whether these findings reflect a true causal effect of blood pressure—potentially through microvascular damage which results in insulin resistance^69^—or whether they are explained by pleiotropic mechanisms influencing both blood pressure and glucose regulation, which is somewhat supported by the attenuated effect estimates from some of the MR methods that are more robust to horizontal pleiotropy.

Overall, our findings were more precisely estimated for SBP than DBP. Given that the selected genetic variants explain a similar amount of variance for SBP and DBP throughout pregnancy, these differences in precision cannot be explained by variation in the strength of the SBP and DBP instruments. We speculate that this might reflect different mechanisms by which higher SBP and DBP impact pregnancy outcomes. For example, high SBP predominantly causes microvascular damage and vessel/cardiac remodeling.^70^ Conversely, elevated DBP may be more indicative of underlying vascular pathology rather than a direct cause of vascular damage and adverse outcomes.^70^Additionally, DBP and SBP are influenced by different physiological mechanisms: DBP reflects a combination of vascular resistance and arterial compliance, which exert opposing effects on its levels.^70^

Our study strengthens the existing evidence base by addressing key limitations of conventional observational studies, which are often prone to confounding, and RCTs, which typically lack sufficient power to detect effects on many clinical outcomes. Unlike traditional observational designs, our MR approach uses genetic variants as instrumental variables, reducing susceptibility to confounding and reverse causation. Furthermore, the use of large-scale datasets provides the required statistical power to detect even modest effects across a broad range of clinical outcomes.

We acknowledge several study limitations. First, the validity of MR findings depends on key assumptions: instrument relevance, independence, and exclusion restriction. Through extensive sensitivity analyses, we demonstrate that the selected SNPs are relevant instruments for the target population, as they are associated with SBP/DBP during pregnancy, as well as HDP, GH, and PE. Moreover, our sensitivity analyses did not indicate that our main findings were substantially biased by population structure or horizontal pleiotropy, including effects from pleiotropic SNPs or fetal genetic contributions. Second, caution is warranted when translating our MR findings into clinical interventions aimed at reducing adverse pregnancy and perinatal outcomes via blood pressure control during pregnancy. This is because the SNPs used cannot accurately estimate the magnitude of blood pressure changes and will predict lifetime variations in blood pressure —spanning preconception, gestation, and post-pregnancy—and therefore cannot inform the optimal dose/timing for therapeutic intervention. Furthermore, interpreting the magnitude of MR effect estimates requires an additional, often unverifiable, assumptions, such as the monotonicity assumption, which posits that the genetic instruments influence the exposure in the same direction across all individuals. Under this assumption we can interpret the results as the average causal effect among those people whose exposure was changed by the genetic instrument, a group that is not necessarily well defined. Finally, we are assuming linear effects of blood pressure on the outcomes, and our sample was mostly restricted to European ancestry women of low-risk pregnancies. Therefore, further evidence is needed to assess transportability to other populations.

## CONCLUSION

In conclusion, our findings suggest that lowering maternal blood pressure is likely to have widespread benefits for maternal and offspring health, including lowering the risk of impaired fetal growth, PTB (including spontaneous PTB), need for induction of labour, GDM and NICU admission. Additionally, our findings indicate that targeting maternal blood pressure is unlikely to affect the risk of miscarriage and stillbirth. Our evidence for perinatal depression and low Apgar at 1 and 5 minutes was less certain due to the modest magnitude and relatively high imprecision of the effect estimates. Overall, our results suggest the need to monitor and manage blood pressure during pregnancy, even in low-risk populations.

## Data Availability

ALSPAC, MoBa, BiB and UK Biobank data are available upon request. ALSPAC data were accessed under project number B3844. The ALSPAC access policy that describes the proposal process in detail including any costs associated with conducting research at ALSPAC, which may be updated from time to time and is available at: https://www.bristol.ac.uk/medialibrary/sites/alspac/documents/researchers/dataaccess/ALSPAC_Access_Policy.pd
Data from the Norwegian Mother, Father and Child Cohort Study and the Medical Birth Registry of Norway used in this study are managed by the national health register holders in Norway (Norwegian Institute of public health) and can be made available to researchers, provided approval from the Regional Committees for Medical and Health Research Ethics (REC), compliance with the EU General Data Protection Regulation (GDPR) and approval from the data owners. The consent given by the participants does not open for storage of data on an individual level in repositories or journals. Researchers who want access to data sets for replication should apply through helsedata.no. Access to data sets requires approval from The Regional Committee for Medical and Health Research Ethics in Norway and an agreement with MoBa (see its website https://www.fhi.no/en/op/data-access-from-health-registries-health-studies-and-biobanks/data-access/applying-for-access-to-data/ for details). Data is available upon request from Born in Bradford: https://borninbradford.nhs.uk/research/how-to-access-data/
Researchers can apply for access to the UK Biobank data via the Access Management System (AMS) (https://www.ukbiobank.ac.uk/enable-your-research/apply-for-access).
Blood pressure GWAS data are publicly available and can be downloaded from the GWAS Catalog (https://www.ebi.ac.uk/gwas/). Statistical code used for the two-sample MR analyses is publicly accessible as part of the 'TwoSampleMR' R package: https://mrcieu.github.io/TwoSampleMR/index.html. The analytic code will be made publicly available upon publication at https://github.com/fernandam93/BP_pregnancy_MR.
This is an Open Access article distributed in accordance with the terms of the Creative Commons Attribution (CC BY 4.0) license, which permits others to distribute, remix, adapt and build upon this work, for commercial use, provided the original work is properly cited. See: http://creativecommons.org/licenses/by/4.0/.

## DECLARATIONS

### Ethics approval and consent to participate

Ethics approval is detailed in the Supplementary methods.

### Consent for publication

Not applicable

Availability of data and materials

ALSPAC, MoBa, BiB and UK Biobank data are available upon request. ALSPAC data were accessed under project number B3844. The ALSPAC access policy that describes the proposal process in detail including any costs associated with conducting research at ALSPAC, which may be updated from time to time and is available at: https://www.bristol.ac.uk/medialibrary/sites/alspac/documents/researchers/dataaccess/ALSPAC_Access_Policy.pd

Data from the Norwegian Mother, Father and Child Cohort Study and the Medical Birth Registry of Norway used in this study are managed by the national health register holders in Norway (Norwegian Institute of public health) and can be made available to researchers, provided approval from the Regional Committees for Medical and Health Research Ethics (REC), compliance with the EU General Data Protection Regulation (GDPR) and approval from the data owners. The consent given by the participants does not open for storage of data on an individual level in repositories or journals. Researchers who want access to data sets for replication should apply through helsedata.no. Access to data sets requires approval from The Regional Committee for Medical and Health Research Ethics in Norway and an agreement with MoBa (see its website https://www.fhi.no/en/op/data-access-from-health-registries-health-studies-and-biobanks/data-access/applying-for-access-to-data/ for details). Data is available upon request from Born in Bradford: https://borninbradford.nhs.uk/research/how-to-access-data/ Researchers can apply for access to the UK Biobank data via the Access Management System (AMS) (https://www.ukbiobank.ac.uk/enable-your-research/apply-for-access). Blood pressure GWAS data are publicly available and can be downloaded from the GWAS Catalog (https://www.ebi.ac.uk/gwas/). Statistical code used for the two-sample MR analyses is publicly accessible as part of the “TwoSampleMR” R package: https://mrcieu.github.io/TwoSampleMR/index.html. The analytic code will be made publicly available upon publication at https://github.com/fernandam93/BP_pregnancy_MR.

This is an Open Access article distributed in accordance with the terms of the Creative Commons Attribution (CC BY 4.0) license, which permits others to distribute, remix, adapt and build upon this work, for commercial use, provided the original work is properly cited. See: http://creativecommons.org/licenses/by/4.0/.

### Competing interests

All authors have completed the ICMJE uniform disclosure form at www.icmje.org/coi_disclosure.pdf and declare: no support from any organisation for the submitted work; no financial relationships with any organisations that might have an interest in the submitted work in the previous three years; no other relationships or activities that could appear to have influenced the submitted work.

## Funding

All cohort specific funding information is detailed in the Supplementary methods.

FMB was funded by a Wellcome Trust four-year PhD studentship (Grant ref: 224982/Z/22/Z). MCB, GLC, DAL, ES, AFS, AGS, QY, FCK, TB, NM and MAA are members of the MRC Integrative Epidemiology Unit at the University of Bristol funded by the UK Medical Research Council (Grant ref: MC_UU_00032/1 and MC_UU_00032/5). QY is also supported by the Noncommunicable Chronic Disease-National Science and Technology Major Project (2024ZD0531500, 2024ZD0531502, 2024ZD0531504). AFS and NM were also supported by Bristol British Heart Foundation (BHF) Accelerator Award (AA/18/7/34219). GLC was additionally supported by the European Union’s Horizon 2020 research and innovation programme (Lifecycle grant 733206). AGS is supported by the European Union’s Horizon 2020 research and innovation programme (874739 LongITools). AGS, GLC and DAL are supported by the European Union’s Horizon Europe Research and Innovation Programme (grant agreement No 101137146, STAGE, via UKRI grant number 10099041). MCB was supported by a Vice-Chancellor’s Research Fellow at the University of Bristol. DAL also received funding from the MRC (CH/F/20/90003) and the NIHR (NF-0616-10102). MCM is supported by the Research Council of Norway through its centers of excellence funding scheme (project number 262700). The funders were not involved in the design of the study, collection and analysis of the data, interpretation of the results, or preparation and submission of the manuscript for publication.

### Authors’ contributions

MCB and GLC contributed equally to this paper and are joint last authors. MCB had the idea for the study and obtained the genetic data. FMB, GLC, and MCB contributed to the study design. AGS, QY, NM, TB, MAA, AFS, GLC, and MCB derived the data for perinatal outcomes. FMB conducted the two-sample MR analysis and produced the tables and figures. All authors contributed to the interpretation of the results. FMB wrote the first draft of the manuscript with the support of AGS, GLC and MCB. All authors revised and approved the final version. FMB, GLC and MCB are the guarantors and attest that all listed authors meet authorship criteria and that no others meeting the criteria have been omitted.

## Acknowledgements

We thank Richard Wilkinson for proofreading several versions of the manuscript. All cohort specific acknowledgements are detailed in the Supplementary methods.

## Transparency

The lead authors affirms that this manuscript is an honest, accurate, and transparent account of the study being reported; that no important aspects of the study have been omitted; and that any discrepancies from the study as planned (and, if relevant, registered) have been explained.

## SUPPLEMENTARY MATERIAL

**Supplement 1.** STROBE-MR checklist of recommended items to address in reports of *Mendelian randomization studies*^25^.

|  | Item N° | Recommendation | Comments/page N° |
| --- | --- | --- | --- |
| **Title and abstract** |  |  |  |
|  | 1 | Indicate Mendelian randomization (MR) as the study's design in the title and/or the abstract if that is a main purpose of the study. | 'Mendelian randomization' in title and abstract |
| **Introduction** |  |  |  |
| Background | 2 | Explain the scientific background and rationale for the reported study. Is a potential causal relationship between exposure and outcome plausible? Justify why MR is a helpful method to address the study question. | Pages 3-4 |
| Objectives | 3 | State specific objectives clearly, including pre-specified causal hypotheses (if any). State that MR is a method that, under specific assumptions, intends to estimate causal effects. | Page 4 |
| **Methods** |  |  |  |
| Study design and data sources | 4 | Present key elements of study design early in the article. Consider including a table listing sources of data for all phases of the study. For each data source contributing to the analysis, describe the following:<br>(a) Setting: Describe the study design and the underlying population, if possible. Describe the setting, locations, and relevant dates, including periods of recruitment,<br>exposure, follow-up, and data collection, when available.<br>(b) Participants: Give the eligibility criteria, and the sources and methods of selection of participants. Report the sample size, and whether any power or sample size calculations were carried out prior to the main analysis.<br>c) Explain how the analysed sample size was arrived at.<br>(d) Describe measurement, quality and selection of genetic variants.<br>(e) For each exposure, outcome and other relevant variables, describe methods of assessment and diagnostic criteria for diseases.<br>(f) Provide details of ethics committee approval and participant informed consent, if relevant. | Pages 4-9, Supplementary methods, Table 1, Table 2, Figure 1 |
| Assumptions | 5 | Explicitly state the three core IV assumptions for the main analysis (relevance, independence, and exclusion restriction) as well assumptions for any additional or sensitivity analysis. | Pages 11-13 |
| Statistical methods: main analysis | 6 | Describe statistical methods and statistics used.<br>(a) Describe how quantitative variables were handled in the analyses (i.e., scale, units, model).<br>(b) Describe how genetic variants were handled in the analyses and, if applicable, how their weights were selected.<br>(c) Describe the MR estimator (e.g. two-stage least squares, Wald ratio) and related statistics. Detail the included covariates and, in case of two-sample MR, whether the same covariate set was used for adjustment in the two samples.<br>(d) Explain how missing data were addressed.<br>(e) If applicable, say how multiple testing was addressed. | Page 9-11, Supplementary methods |
| Assessment of assumptions | 7 | Describe any methods or prior knowledge used to assess the assumptions or justify their validity. | Pages 11-13, Supplementary methods |
| Sensitivity analyses | 8 | Describe any sensitivity analyses or additional analyses performed (e.g. comparison of effect estimates from different approaches, independent replication, bias analytic techniques, validation of instruments, simulations). | Pages 11-13, Supplementary methods |
| Software and pre-registration | 9 | (a) Name statistical software and package(s), including version and settings used.<br>(b) State whether the study protocol and details were pre-registered (as well as when and where). | Page 14, Supplementary methods |
| **Results** |  |  |  |
| Descriptive data | 10 | (a) Report the numbers of individuals at each stage of included studies and reasons for exclusion. Consider use of a flow diagram.<br>(b) Report summary statistics for phenotypic exposure(s), outcome(s) and other relevant variables (e.g. means, SDs, proportions).<br>(c) If the data sources include meta-analyses of previous studies, provide the assessments of heterogeneity across these studies.<br>(d) For two-sample MR:<br>i. Provide justification of the similarity of the genetic variant-exposure associations between the exposure and<br>outcome samples.<br>*ii.* Provide information on the number of individuals who overlap between the exposure and outcome studies. | Not applicable, as we used summary statistics from previously published genome-wide association studies. We cite these accordingly. Sample overlap: page 7 |
| Main results | 11 | (a) Report the associations between genetic variant and exposure, and between genetic variant and outcome, preferably on an interpretable scale.<br>(b) Report MR estimates of the relationship between exposure and outcome, and the measures of uncertainty from the MR analysis on an interpretable scale, such as odds ratio or relative risk per SD difference.<br>(c) If relevant, consider translating estimates of relative risk into absolute risk for a meaningful time-period.<br>(d) Consider plots to visualize results (e.g. forest plot, scatterplot of associations between genetic variants and outcome versus between genetic variants and exposure). | Pages 14-20, Figures 2-5, Supplementary figures and tables |
| Assessment of validity | 12 | (a) Report the assessment of the validity of the assumptions.<br>(b) Report any additional statistics (e.g., assessments of heterogeneity across genetic variants, such as $I^2$, $Q$ statistic or $E$-value). | Pages 14-20 and Figures 3-5, Supplementary figures and tables |
| Sensitivity and additional analyses | 13 | (a) Report any sensitivity analyses to assess the robustness of the main results to violations of the assumptions.<br>(b) Report results from other sensitivity analyses or additional analyses.<br>(c) Report any assessment of direction of causal relationship (e.g., bidirectional MR).<br>(d) When relevant, report and compare with estimates from non-MR analyses.<br>(e) Consider any additional plots to visualize results (e.g., leave-one-out analyses). | Pages 14-20 and Figures 3-5, Supplementary figures and tables |
| **Discussion** |  |  |  |
| Key results | 14 | Summarize key results with reference to study objectives. | Page 21 |
| Limitations | 15 | Discuss limitations of the study, taking into account the validity of the IV assumptions, other sources of potential bias, and imprecision. Discuss both direction and magnitude of any potential bias, and any efforts to address them. | Pages 21-24 |
| Interpretation | 16 | (a) Meaning: Give a cautious overall interpretation of results in the context of their limitations and in comparison with other studies.<br>(b) Mechanisms: Discuss underlying biological mechanisms that could drive a potential causal relationship between the investigated exposure and the outcome, and whether the gene-environment equivalence assumption is reasonable. Use causal language carefully, clarifying that IV estimates may provide causal effects only under certain assumptions.<br>(c) Clinical relevance: Discuss whether the results have clinical or public policy relevance, and to what extent they inform effect sizes of possible interventions. | Pages 21-24 |
| Generalisability | 17 | Discuss the generalizability of the study results (a) to other populations (i.e. external validity), (b) across other exposure periods/timings, and (c) across other levels of exposure. | Page 24 |
| **Other information** |  |  |  |
| Funding | 18 | Describe sources of funding and the role of funders in the present study and, if applicable, sources of funding for the databases and original study or studies on which the present study is based. | Pages 31-32 |
| Data and data sharing | 19 | Provide the data used to perform all analyses or report where and how the data can be accessed, and reference these sources in the article. Provide the statistical code needed to reproduce the results in the article, or report whether the code is publicly accessible and if so, where. | Pages 30-31, Supplementary methods |
| Conflicts of interest | 20 | All authors should declare all potential conflicts of interest. | Page 31 |

Adapted from: BMJ 2021;375:n2233 http://dx.doi.org/10.1136/bmj.n2233

## Notes

### Competing Interest Statement

The authors have declared no competing interest.

### Author Declarations

ALSPAC Ethical approval was obtained from the ALSPAC Ethics and Law Committee and the Local Research Ethics Committees. Consent for biological samples has been collected in accordance with the Human Tissue Act (2004). Informed consent for the use of data collected via questionnaires and clinics was obtained from participants following the recommendations of the ALSPAC Ethics and Law Committee at the time (details and reference numbers of all ethics approvals can be found at http://www.bristol.ac.uk/media-library/sites/alspac/documents/governance/Research%20Ethics%20Committee%20approval%20references.pdf). BiB Ethical approval for the study was granted by the Bradford National Health Service Research Ethics Committee (ref 06/Q1202/48), and all participants gave written informed consent. The ALL IN sub-study had ethical approval from the London School of Hygiene & Tropical Medicine ethics committee (ref: 5320) and the Bradford Research Ethics committee (ref: 08/H1302/21). Parents (usually the mother) gave informed, written consent to take part in the study. MoBa The current study is based on version 12 of the quality-assured data files released for research in 2019. The establishment of MoBa and initial data collection was based on a license from the Norwegian Data Protection Agency and approval from The Regional Committees for Medical and Health Research Ethics. The MoBa cohort is currently regulated by the Norwegian Health Registry Act. The current study was approved by The Regional Committees for Medical and Health Research Ethics of South/East Norway (ref 2018/1256). UK Biobank The UK Biobank has approval from the North West Multi-centre Research Ethics Committee (MREC) as a Research Tissue Bank (RTB) approval. This RTB approval was granted initially in 2011 (11/NW/0382) and it is renewed on a 5-yearly cycle, with the latest one successfully renewed in 2021 (21/NW/0157).

### Summary of Updates

In the methods section, we added the methods for the positive control outcome analysis and edited the flowchart to only include positive control outcomes.

