## Supplementary figures for "Assessing the impact of maternal blood pressure during pregnancy on perinatal health: A wide-angled Mendelian randomization study"

**Supplementary Figures 1a-e**. Correlation plots for SBP/DBP instrumental variables and maternal blood pressure during pregnancy (i.e., first, second and third trimesters and across gestation), GH, preeclampsia and HDP.

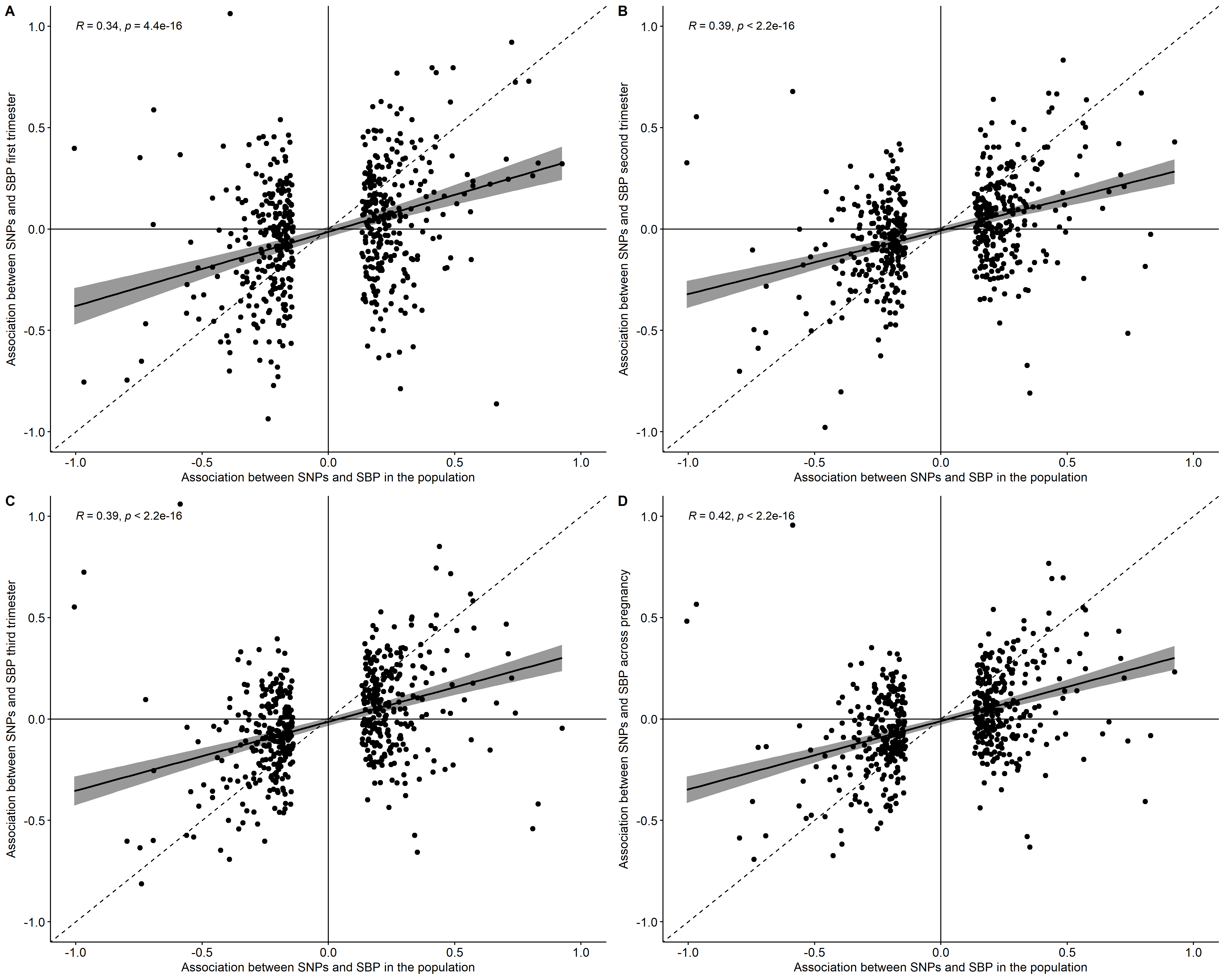

### Supplementary Figure 1a (Correlation for SBP IVs and SBP during pregnancy)

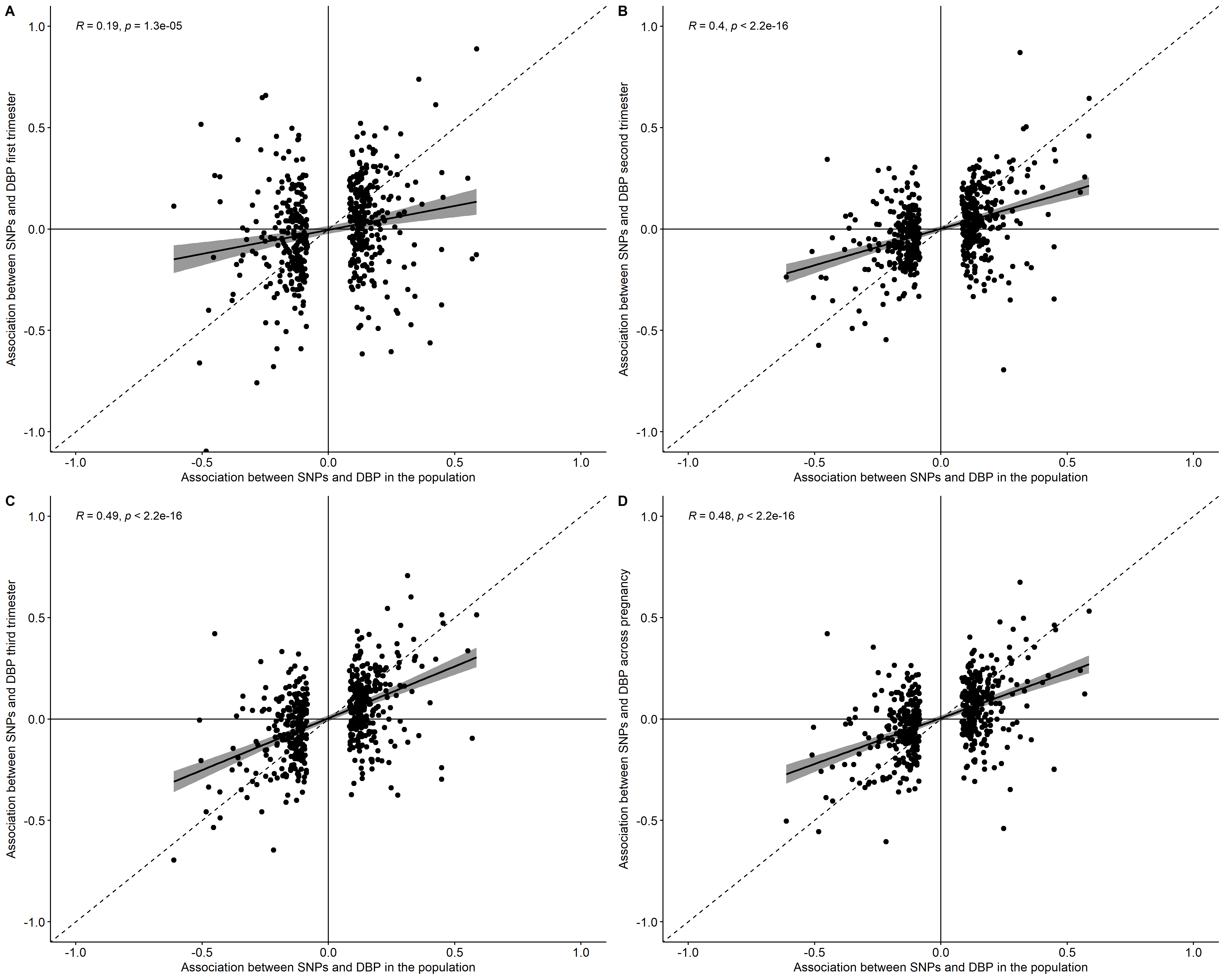

### Supplementary Figure 1b (Correlation for DBP IVs and DBP during pregnancy)

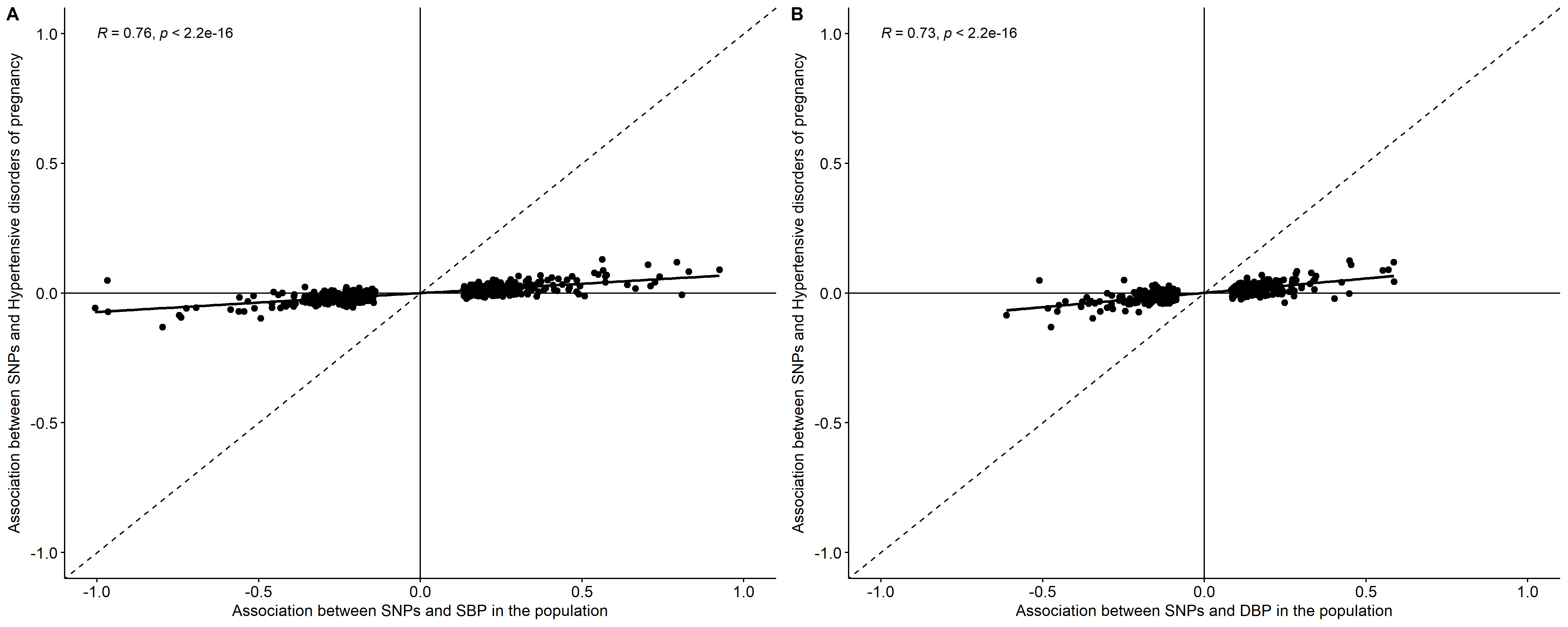

### Supplementary Figure 1c (Correlation for SBP/DBP IVs and HDP)

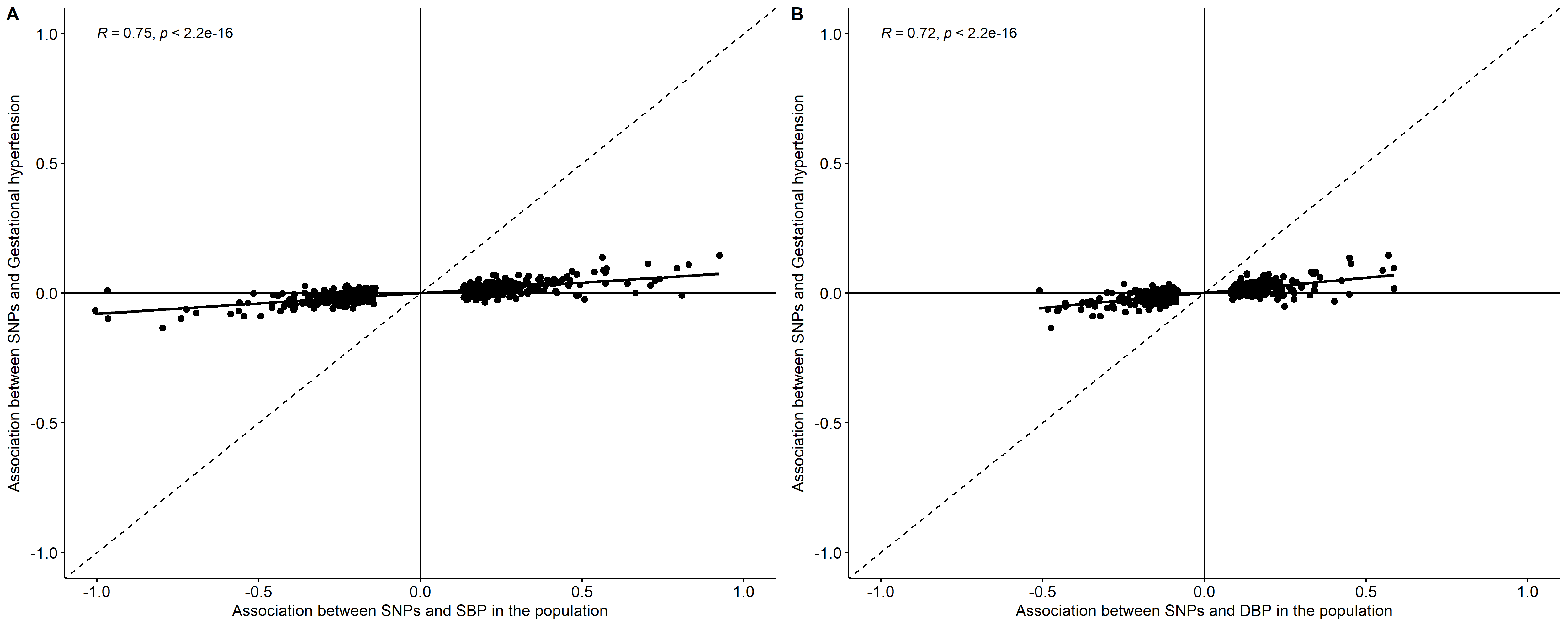

### Supplementary Figure 1d (Correlation for SBP/DBP IVs and GH)

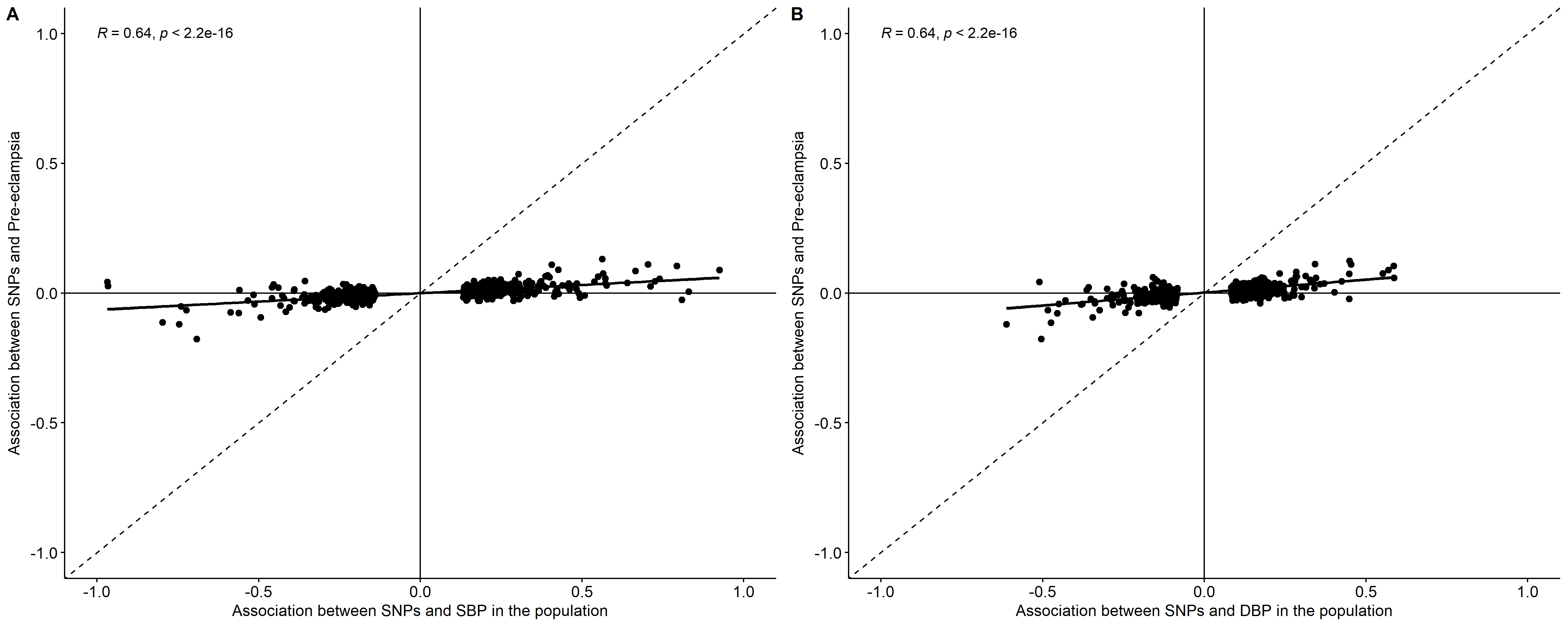

### Supplementary Figure 1e (Correlation for SBP/DBP IVs and PE)

**Supplementary Figures 2a-e**. Regression plots for SBP/DBP instrumental variables and maternal blood pressure during pregnancy (i.e., first, second and third trimesters and across gestation), GH, preeclampsia and HDP.

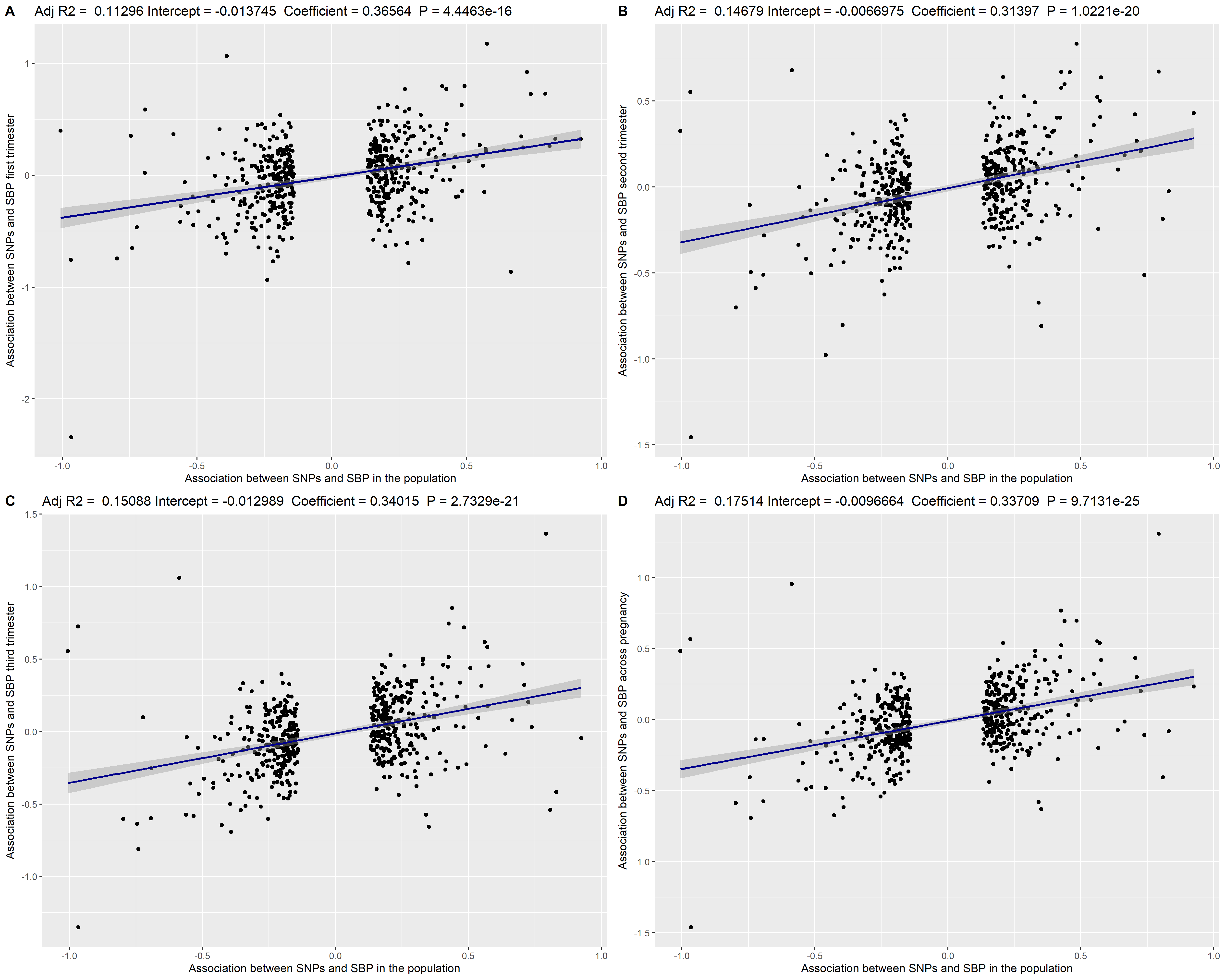

### Supplementary Figure 2a (Regression for SBP IVs and SBP during pregnancy)

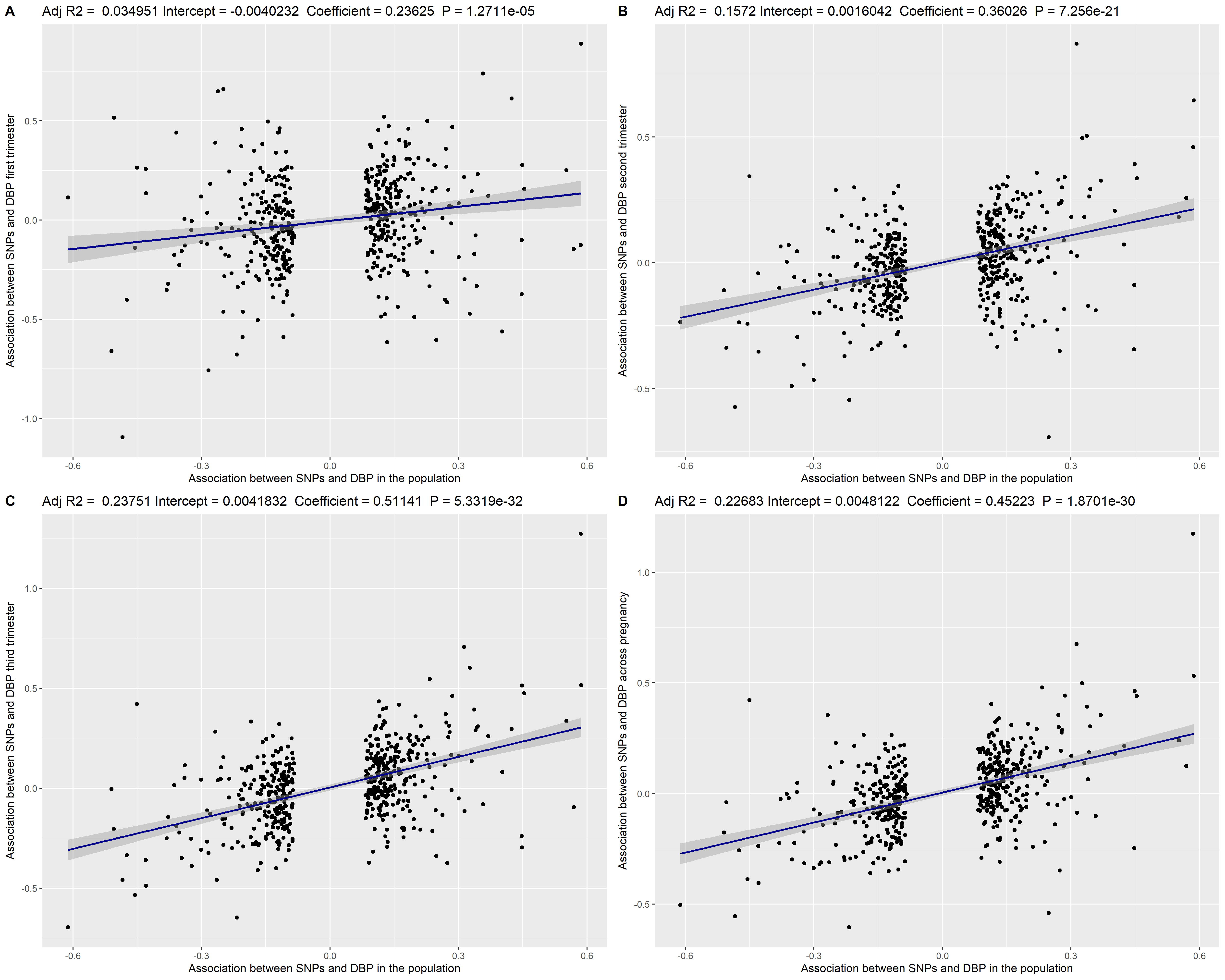

### Supplementary Figure 2b (Regression for DBP IVs and DBP during pregnancy)

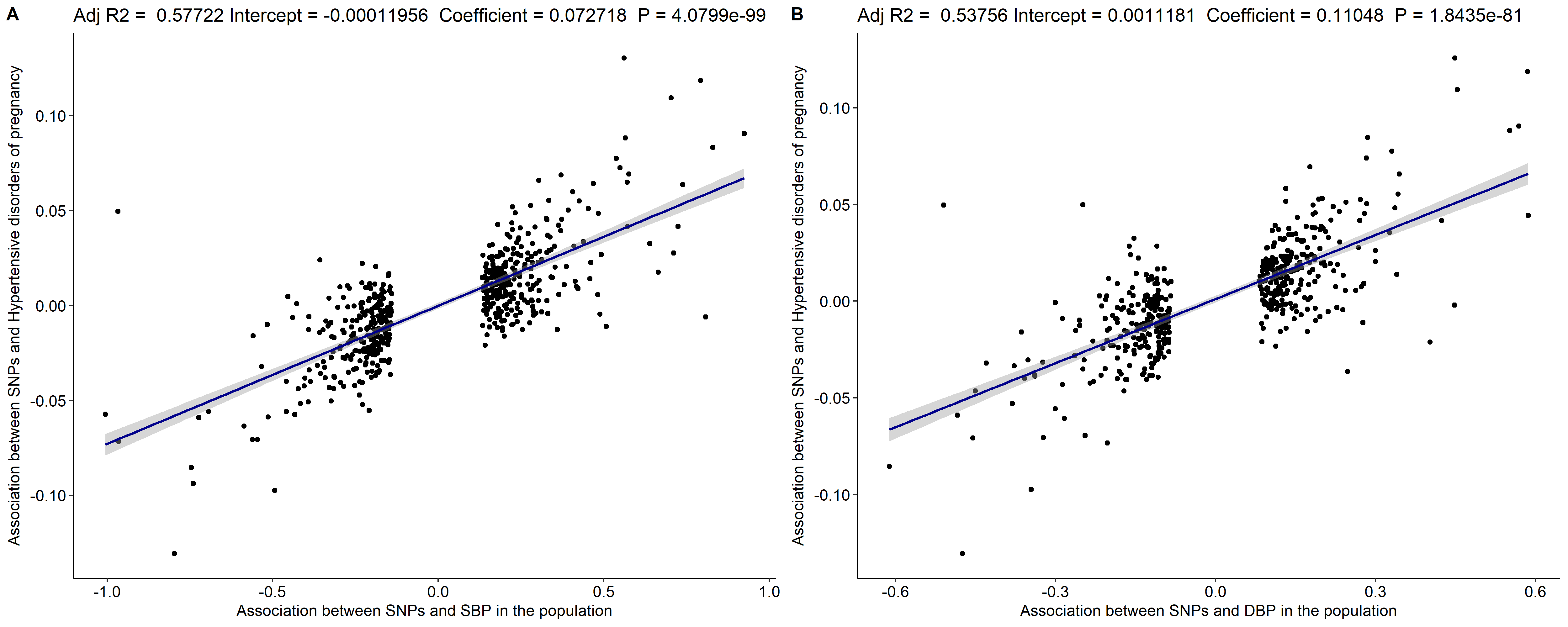

### Supplementary Figure 2c (Regression for SBP/DBP IVs and HDP)

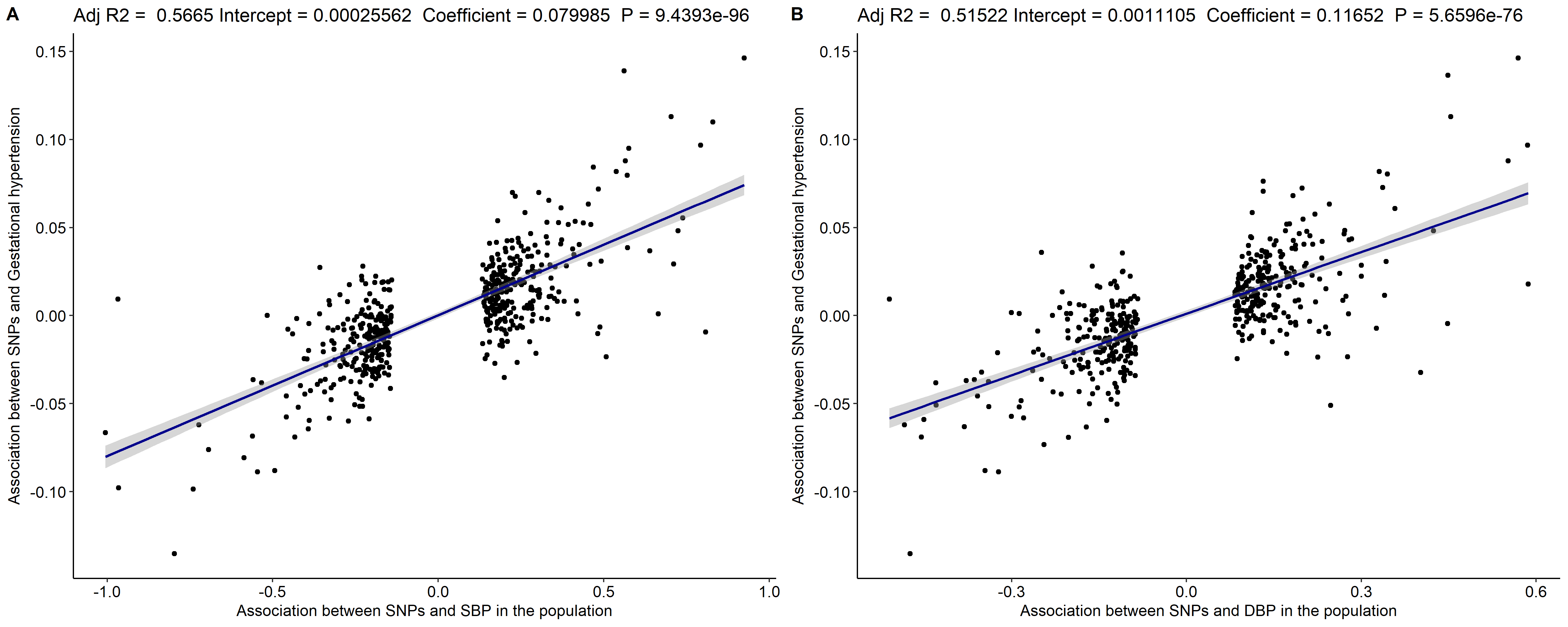

### Supplementary Figure 2d (Regression for SBP/DBP IVs and GH)

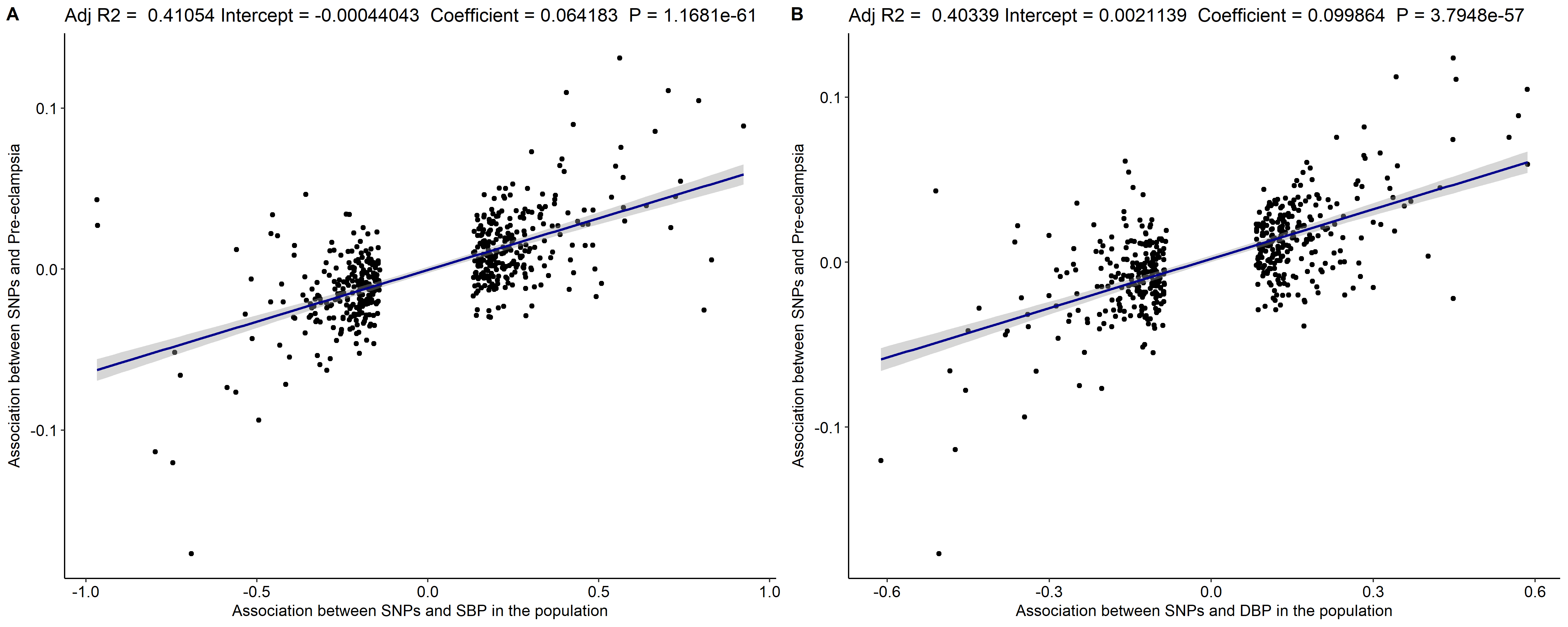
Supplementary Figure 2e (Regression for SBP/DBP IVs and PE)

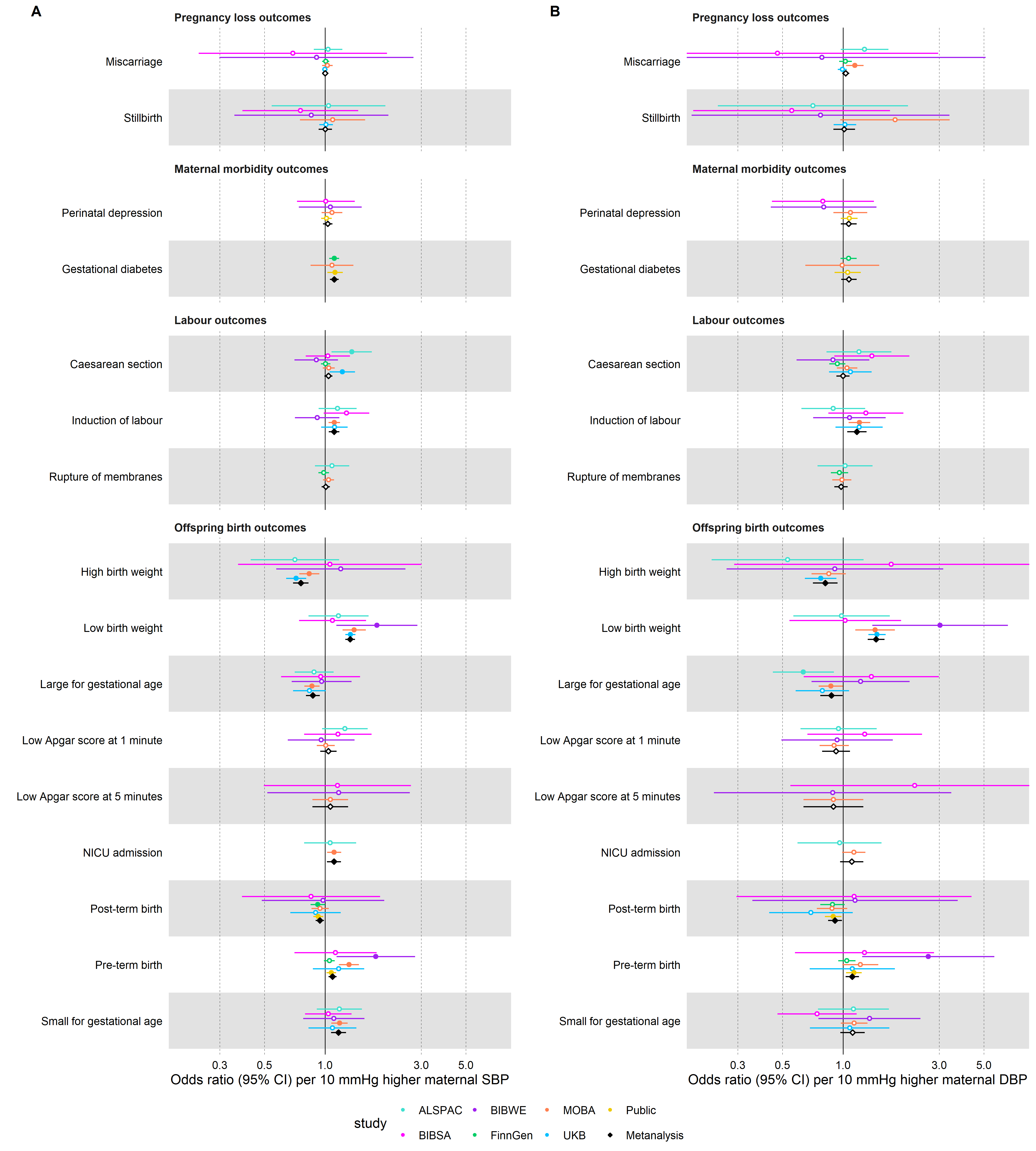

### **Supplementary Figure 3**. Study-specific and pooled Mendelian randomization estimates for the genetically predicted effects of maternal blood pressure on the primary outcomes.

Odds ratios and 95% confidence intervals are presented per 10 mmHg increase in (A) systolic and (B) diastolic blood pressure.

Systolic and diastolic blood pressure were instrumented by 545 and 513 genetic variants obtained from Keaton et al.^45^, respectively.

Abbreviations: SBP, systolic blood pressure; DBP, diastolic blood pressure.

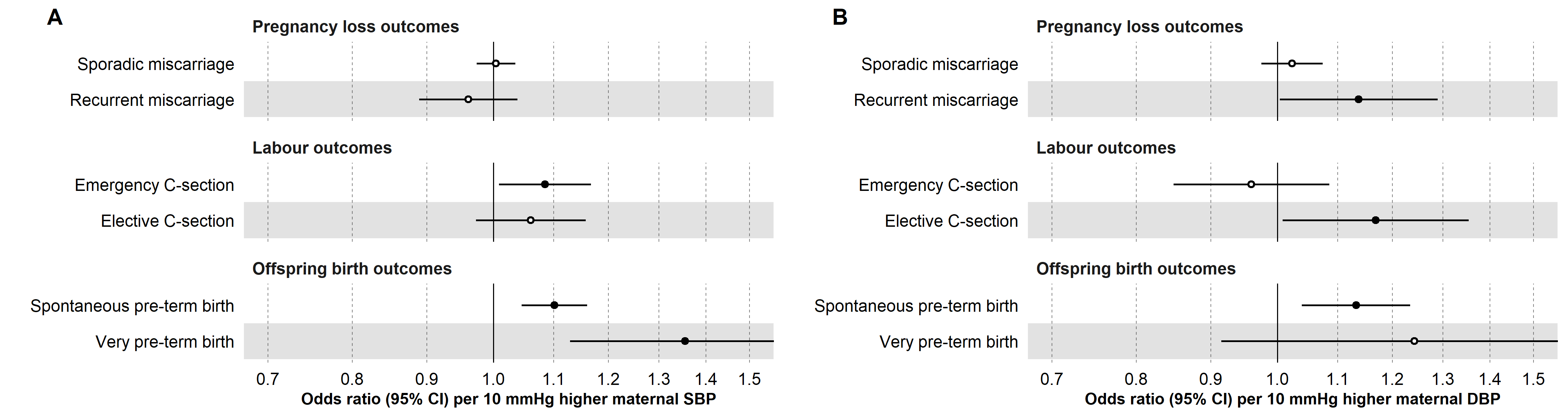

### **Supplementary Figure 4**. Inverse-variance weighted Mendelian randomization estimates for genetically predicted effects of maternal blood pressure on the secondary (binary) outcomes.

Odds ratios and 95% confidence intervals are presented per 10 mmHg increase in (A) systolic and (B) diastolic blood pressure.

Systolic and diastolic blood pressure were instrumented by 545 and 513 genetic variants obtained from Keaton et al.^45^, respectively.

Abbreviations: SBP, systolic blood pressure; DBP, diastolic blood pressure.

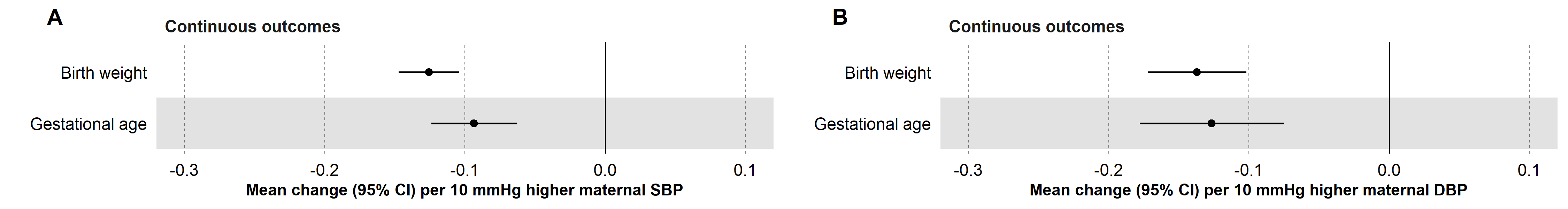

### **Supplementary Figure 5**. Inverse-variance weighted Mendelian randomization estimates for genetically predicted effects of maternal blood pressure on the secondary (continuous) outcomes.

Coefficients and 95% confidence intervals for birthweight (in standard deviation units) and gestational age (in weeks) are presented per 10 mmHg increase in (A) systolic and (B) diastolic blood pressure. Systolic and diastolic blood pressure were instrumented by 545 and 513 genetic variants obtained from Keaton et al.^45^, respectively.

Abbreviations: SBP, systolic blood pressure; DBP, diastolic blood pressure.

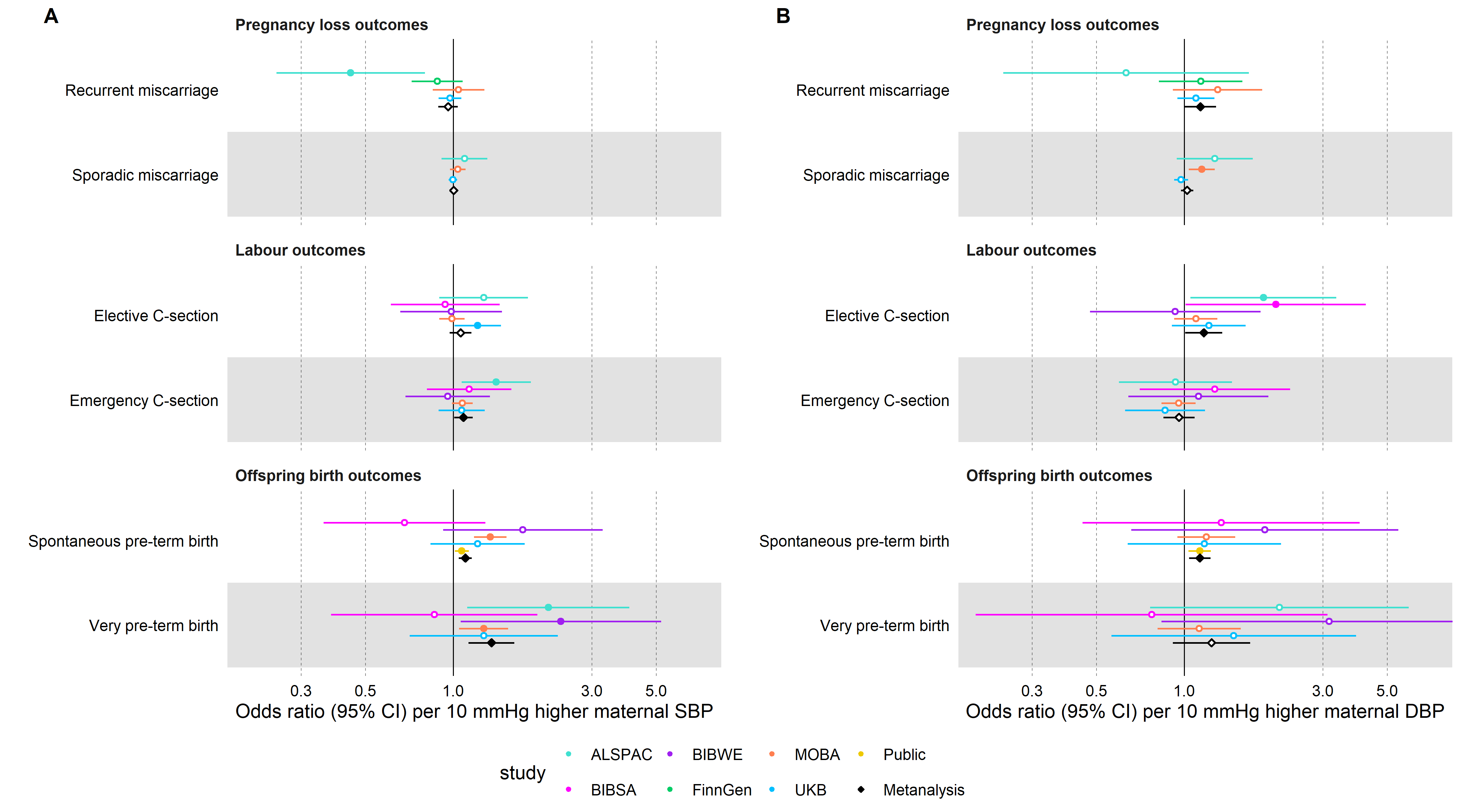

### **Supplementary Figure 6.** Study-specific and pooled Mendelian randomization estimates for the genetically predicted effects of maternal blood pressure on the secondary (binary) outcomes.

Odds ratios and 95% confidence intervals are presented per 10 mmHg increase in (A) systolic and (B) diastolic blood pressure.

Systolic and diastolic blood pressure were instrumented by 545 and 513 genetic variants obtained from Keaton et al.^45^, respectively.

Abbreviations: SBP, systolic blood pressure; DBP, diastolic blood pressure.

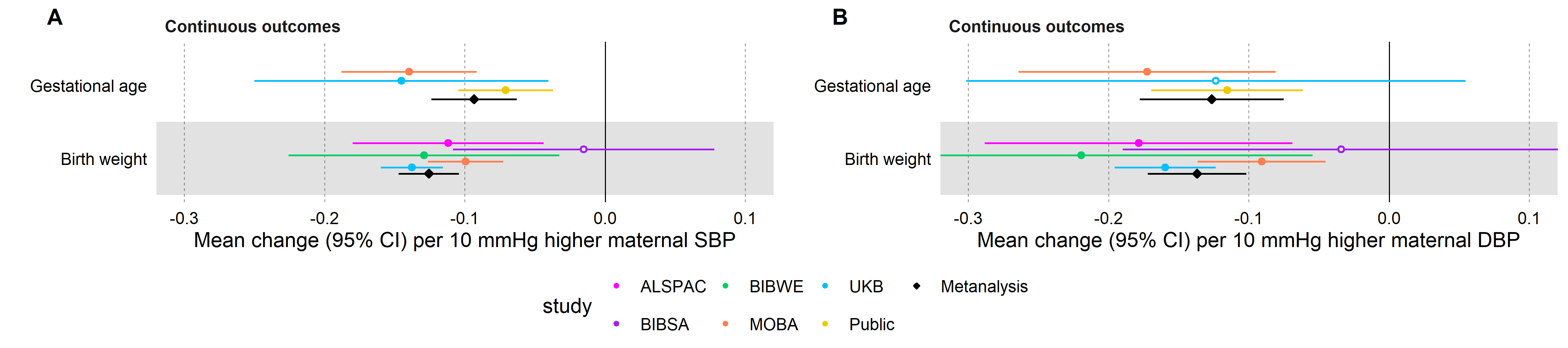

### **Supplementary Figure 7**. Study-specific and pooled Mendelian randomization estimates for the genetically predicted effects of maternal blood pressure on the secondary (continuous) outcomes.

Coefficients and 95% confidence intervals for birthweight (in standard deviation units) and gestational age (in weeks) are presented per 10 mmHg increase in (A) systolic and (B) diastolic blood pressure. Systolic and diastolic blood pressure were instrumented by 545 and 513 genetic variants obtained from Keaton et al.^45^, respectively.

Abbreviations: SBP, systolic blood pressure; DBP, diastolic blood pressure.

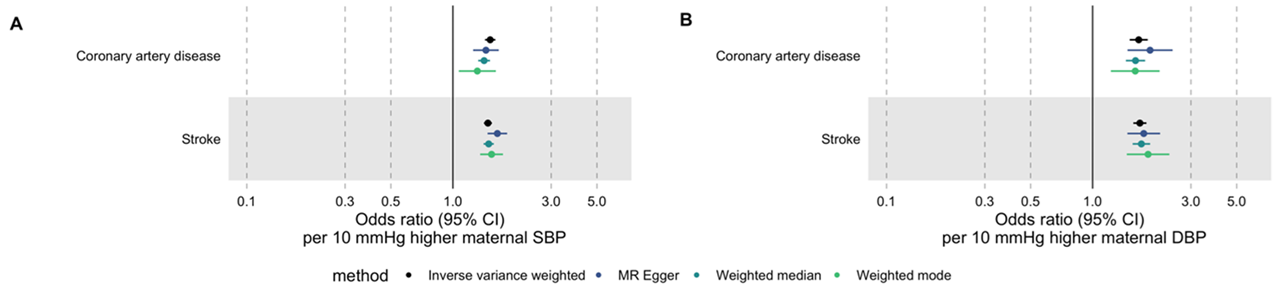

### **Supplementary Figure 8**. Mendelian randomization estimates for genetically predicted effects of maternal blood pressure on positive control outcomes.

Outcomes were obtained using the IEU OpenGWAS API. Dataset IDs for positive control outcomes: stroke (ebi-a-GCST005838) and coronary artery disease (ebi-a-GCST003116).

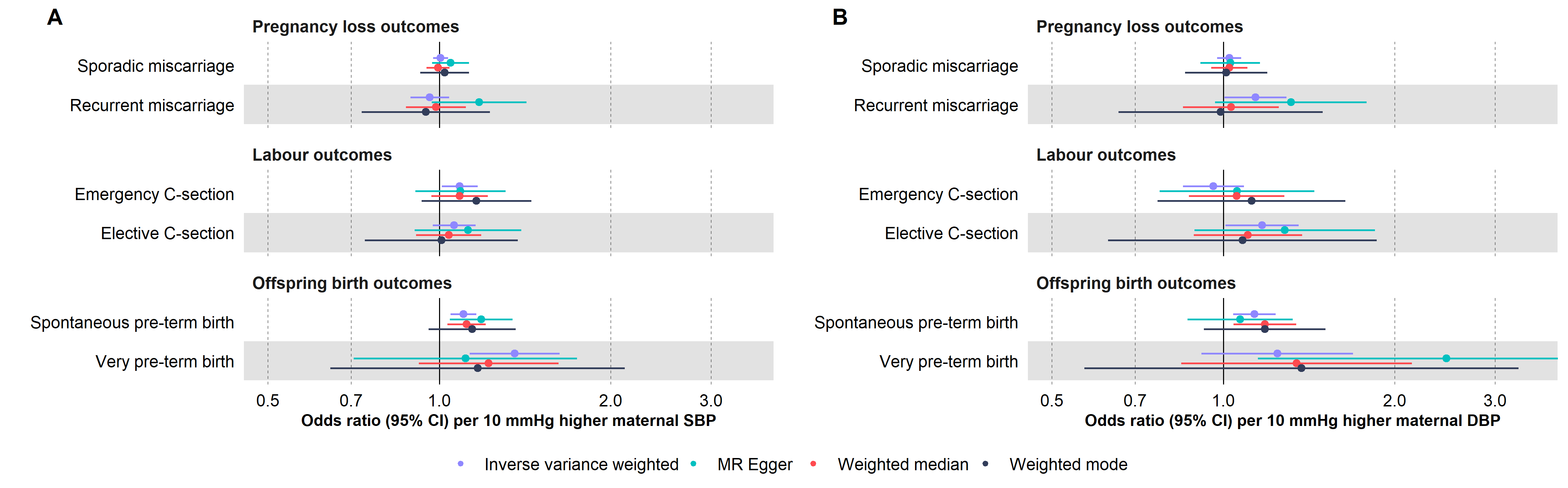

### **Supplementary Figure 9**. Mendelian randomization estimates for genetically predicted effects of maternal blood pressure on the secondary (binary) outcomes across different.

Odds ratios and 95% confidence intervals are presented per 10 mmHg increase in (A) systolic and (B) diastolic blood pressure.

Systolic and diastolic blood pressure were instrumented by 545 and 513 genetic variants obtained from Keaton et al.^45^, respectively.

Abbreviations: SBP, systolic blood pressure; DBP, diastolic blood pressure.

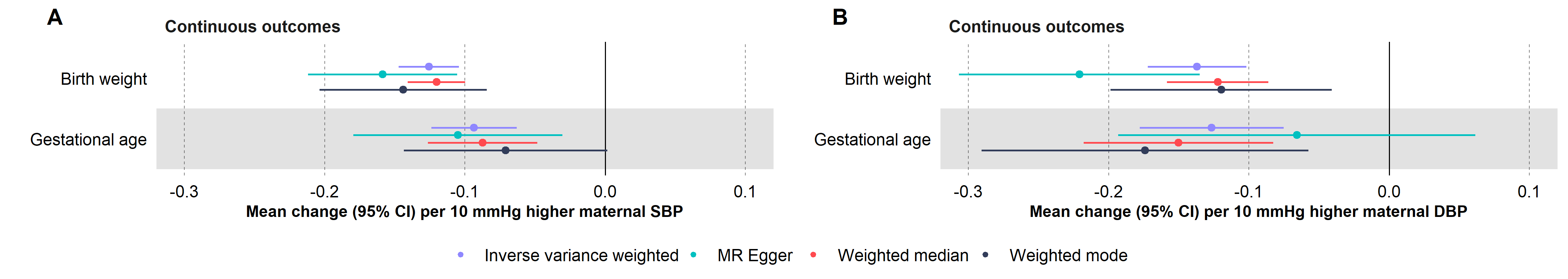

### **Supplementary Figure 10**. Mendelian randomization estimates for genetically predicted effects of maternal blood pressure on the secondary (continuous) outcomes across different methods

Coefficients and 95% confidence intervals for birthweight (in standard deviation units) and gestational age (in weeks) are presented per 10 mmHg increase in (A) systolic and (B) diastolic blood pressure. Systolic and diastolic blood pressure were instrumented by 545 and 513 genetic variants obtained from Keaton et al.^45^, respectively.

Abbreviations: SBP, systolic blood pressure; DBP, diastolic blood pressure.

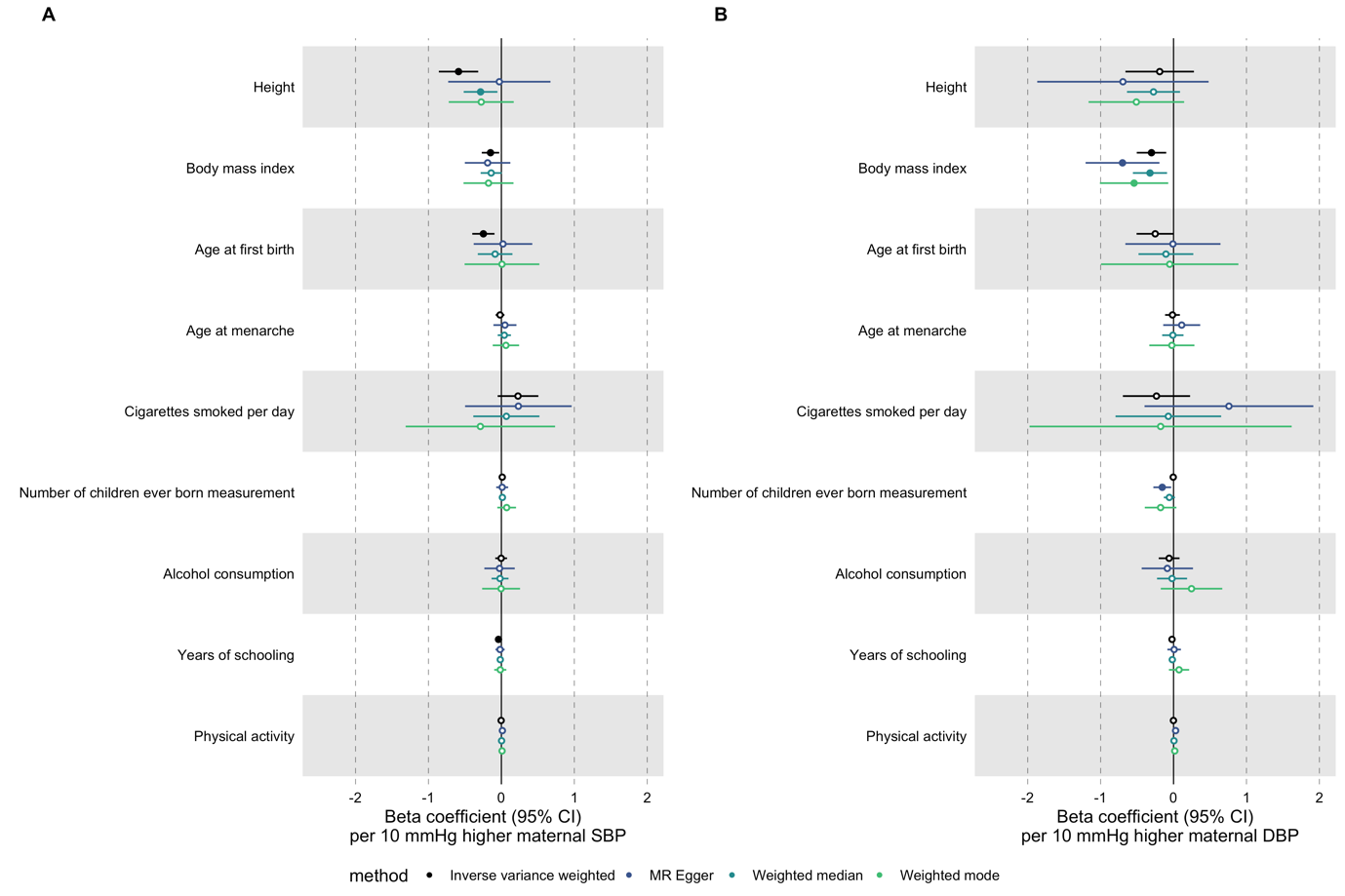

### **Supplementary Figure 11**. Mendelian randomization estimates for genetically predicted effects of maternal blood pressure on determinants of perinatal health

i.e., height (id: ieu-b-4814), body mass index (BMI; id: ieu-b-4816), age at first birth (id: ieu-b-4820), age at menarche (id: ieu-b-4822), cigarettes smoked per day (id: ieu-b-4826), number of children ever born (id: ieu-b-4828), alcohol consumption (id: ieu-b-4834), years of schooling (id: ieu-b-4836) and physical activity (id: ieu-b-4860) in the UK Biobank.

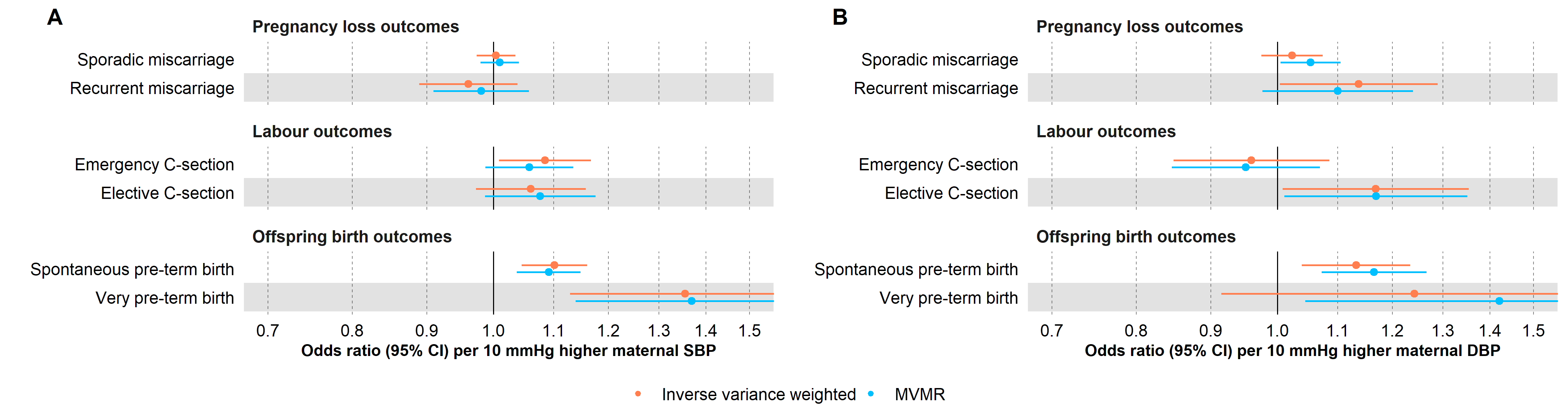

### **Supplementary Figure 12**. Univariable (“Inverse variance weighted”) and multivariable (“MVMR”) Mendelian randomization estimates for genetically predicted effects of maternal blood pressure on the secondary outcomes. Multivariable models have been adjusted for body mass index, height, years of education and age at first birth.

Odds ratios and 95% confidence intervals are presented per 10 mmHg increase in (A) systolic and (B) diastolic blood pressure.

Systolic and diastolic blood pressure were instrumented by 545 and 513 genetic variants obtained from Keaton et al.^45^, respectively.

Abbreviations: SBP, systolic blood pressure; DBP, diastolic blood pressure.

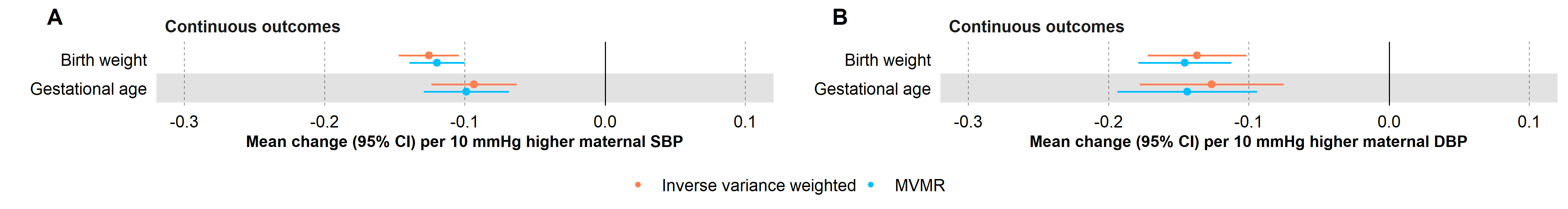

### **Supplementary Figure 13**. Univariable and multivariable Mendelian randomization estimates for genetically predicted effects of maternal blood pressure on binary perinatal outcomes. Multivariable models have been adjusted for body mass index, a potential collider.

Coefficients and 95% confidence intervals for birthweight (in standard deviation units) and gestational age (in weeks) are presented per 10 mmHg increase in (A) systolic and (B) diastolic blood pressure. Systolic and diastolic blood pressure were instrumented by 545 and 513 genetic variants obtained from Keaton et al.^45^, respectively.

Abbreviations: SBP, systolic blood pressure; DBP, diastolic blood pressure.

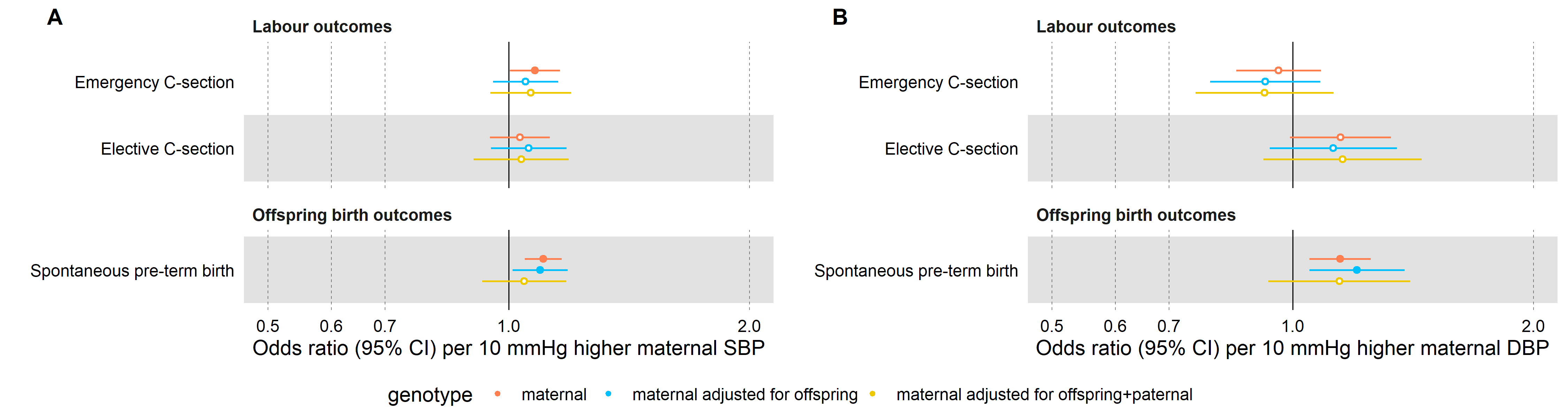

### **Supplementary Figure 14**. Mendelian randomization estimates for genetically predicted effects of maternal blood pressure on the secondary binary outcomes, adjusting for offspring genotype.

Odds ratios and 95% confidence intervals are presented per 10 mmHg increase in (A) systolic and (B) diastolic blood pressure. Estimates are shown for maternal effects unadjusted (red) and adjusted by fetal genetic effects (blue), and adjusted by both offspring and paternal genetic effects (yellow). Systolic and diastolic blood pressure were instrumented by 545 and 513 genetic variants obtained from Keaton et al.^45^, respectively.

Abbreviations: SBP, systolic blood pressure; DBP, diastolic blood pressure.

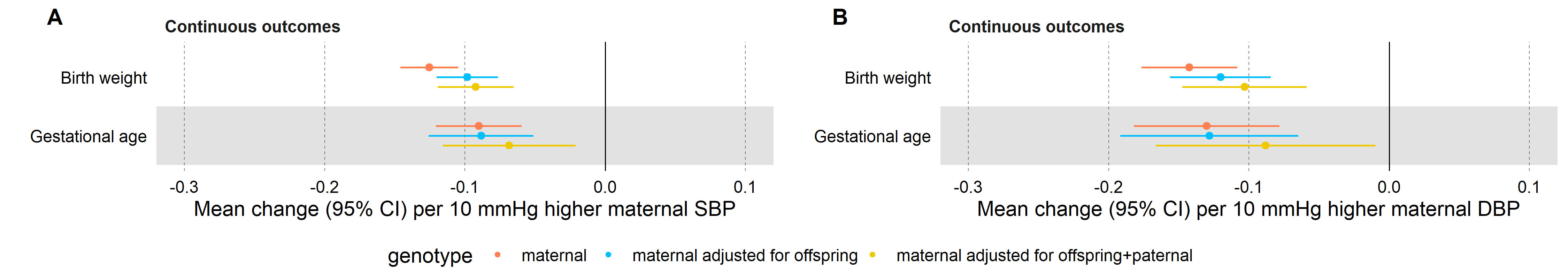

### **Supplementary Figure 15**. Mendelian randomization estimates for genetically predicted effects of maternal blood pressure on the secondary continuous outcomes, adjusting for offspring genotype.

Coefficients and 95% confidence intervals for birthweight (in standard deviation units) and gestational age (in weeks) are presented per 10 mmHg increase in (A) systolic and (B) diastolic blood pressure. Estimates are shown for maternal effects unadjusted (red) and adjusted by fetal genetic effects (blue), and adjusted by both offspring and paternal genetic effects (yellow). Systolic and diastolic blood pressure were instrumented by 545 and 513 genetic variants obtained from Keaton et al.^45^, respectively.

Abbreviations: SBP, systolic blood pressure; DBP, diastolic blood pressure.

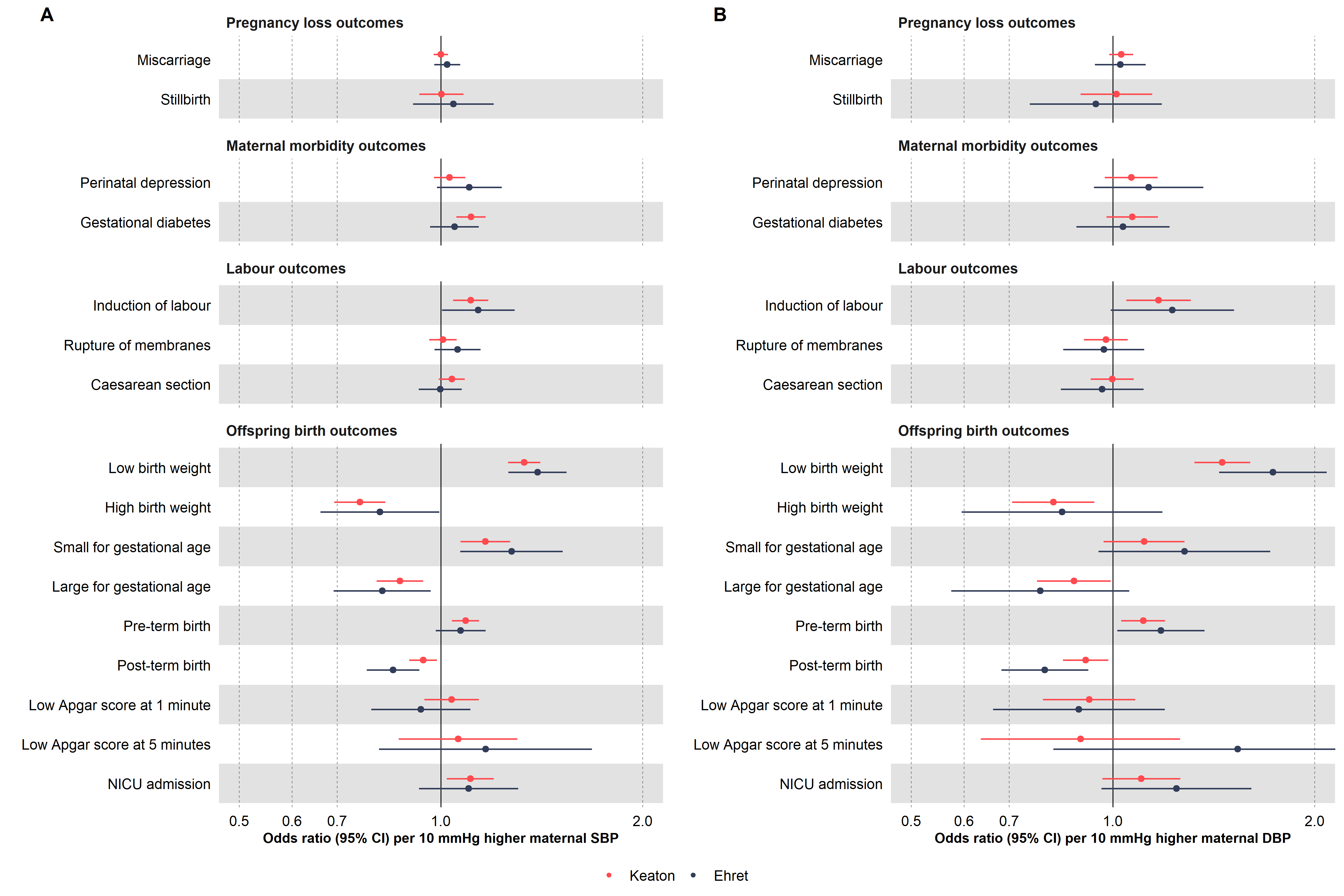

### **Supplementary Figure 16**. Mendelian randomization estimates for genetically predicted effects of maternal blood pressure on the primary outcomes in overlapping (Keaton et al.) and non-overlapping samples (Ehret et al.).

Odds ratios and 95% confidence intervals are presented per 10 mmHg increase in (A) systolic and (B) diastolic blood pressure. Systolic and diastolic blood pressure were instrumented by 545 and 513 genetic variants obtained from Keaton et al.^45^, respectively.

Abbreviations: SBP, systolic blood pressure; DBP, diastolic blood pressure; NICU, neonatal intensive care unit.

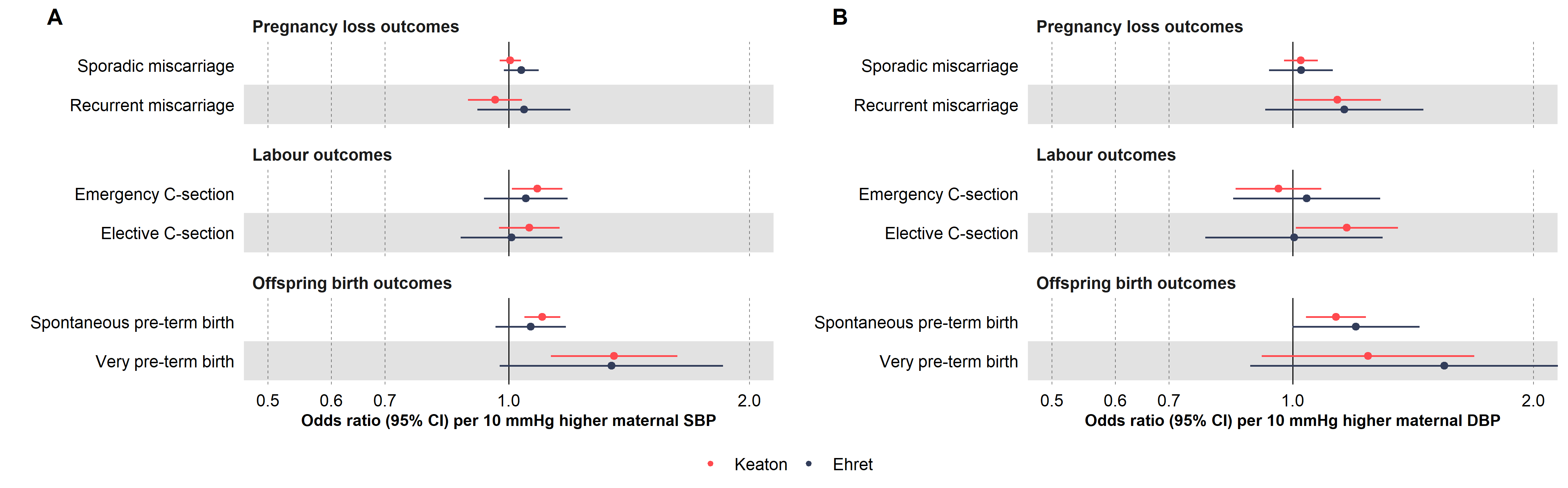

### **Supplementary Figure 17**. Mendelian randomization estimates for genetically predicted effects of maternal blood pressure on the secondary (binary) outcomes in overlapping (Keaton et al.) and non-overlapping samples (Ehret et al.).

Odds ratios and 95% confidence intervals are presented per 10 mmHg increase in (A) systolic and (B) diastolic blood pressure. Systolic and diastolic blood pressure were instrumented by 545 and 513 genetic variants obtained from Keaton et al.^45^, respectively.

Abbreviations: SBP, systolic blood pressure; DBP, diastolic blood pressure.

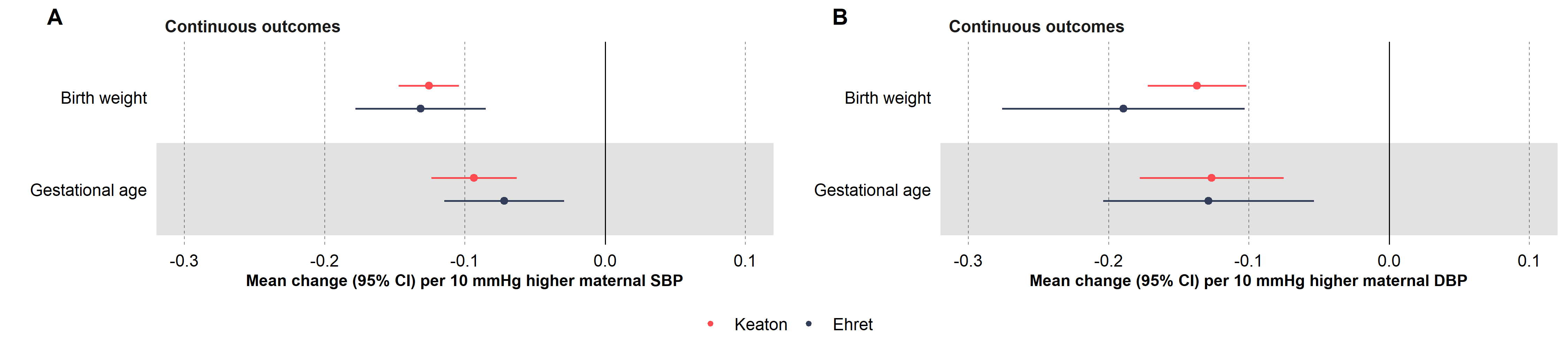

### **Supplementary Figure 18**. Mendelian randomization estimates for genetically predicted effects of maternal blood pressure on the secondary (continuous) outcomes in overlapping (Keaton et al.) and non-overlapping samples (Ehret et al.).

Coefficients and 95% confidence intervals for birthweight (in standard deviation units) and gestational age (in weeks) are presented per 10 mmHg increase in (A) systolic and (B) diastolic blood pressure. Systolic and diastolic blood pressure were instrumented by 545 and 513 genetic variants obtained from Keaton et al.^45^, respectively.

Abbreviations: SBP, systolic blood pressure; DBP, diastolic blood pressure.
