## Supplementary Methods for "Assessing the impact of maternal blood pressure during pregnancy on perinatal health: A wide-angled Mendelian randomization study"

### Data sources

The data sources which are part of the MR-PREG collaboration are described briefly below and in more detail elsewhere^1^.

#### Avon Longitudinal Study of Parents and Children (ALSPAC)

1. **Overview**

ALSPAC is a prospective birth cohort that started recruiting pregnant women resident in the former county of Avon (centred around the city of Bristol), with delivery dates between April 1991 and December 1992. A total of 14,541 women (ALSPAC-G0) were enrolled during pregnancy (14,676 fetuses) and gave birth to 14,062 live children (ALSPAC-G1) ^2 3^. Women responded to four questionnaires during pregnancy (average 8-, 12-, 18-, and 32-weeks’ gestation) and two postpartum (average 8 weeks and 8 months). Biological samples were collected during pregnancy (blood and urine) and birth (cord blood and placenta). Maternal anthropometrics was based on self-report collected at 12 weeks gestation and children had anthropometrics measured at birth. Obstetric records were also linked to the participants. Genetic data is available for mothers, partners and children.

ALSPAC contributed to this study with data on maternal genotype and perinatal outcomes that were used in the main analyses, as well as offspring genotype and blood pressure traits during pregnancy (i.e., SBP, DBP, GH, preeclampsia and HDP) that were used in sensitivity analyses.

2. ***Genetic data***

Maternal genetic data were generated using the Illumina Human660W-Quad genotyping array at the *Centre National de Génotypage* (CNG, Paris, France). Offspring genetic data were generated using the Illumina HumanHap550-Quad genotyping array at the Wellcome Trust Sanger Institute (WTSI, Cambridge, UK) and the Laboratory Corporation of America (LCA, Burlington, NC, US). Imputation was conducted on the Michigan Imputation Server, using the Haplotype Reference Consortium (HRC r1.1) as the reference panel. Quality control filtering was performed using PLINK v1.07 and it excluded individuals based on relatedness, non-European ancestry and gender mismatches.

3. ***Blood pressure traits during pregnancy***

Maternal blood pressure data were obtained from obstetric records. Seated SBP and DBP were taken by obstetricians or midwives as part of routine prenatal medical care. Measurements were not recorded specifically for ALSPAC research. GH was defined as SBP ≥140 mmHg and/or DBP ≥90 mmHg, measured on ≥2 occasions after 20 weeks of gestation in absence of proteinuria (<1+ on urine dipstick), in women who did not report having hypertension before pregnancy. Preeclampsia was defined as SBP ≥140 mmHg and/or DBP ≥90 mmHg, measured on ≥2 occasions after 20 weeks of gestation in presence of proteinuria (≥1+ on urine dipstick), in women who did not report having hypertension before pregnancy. HDP comprised both GH and preeclampsia.

4. ***Perinatal outcomes***

ALSPAC contributed with data for perinatal outcomes as described in **Supplementary table 4**.

5. ***Summary data for genotype-phenotype associations***

Summary data for genotype-phenotype associations were generated using linear/logistic regression, implemented using PLINK 2.0 alpha (build 22 Dec 2024) (13, 14). The analytical samples were restricted to unrelated mothers (IBD ≤ 0.125) and offspring (IBD ≤ 0.1). Association analyses were performed with imputed autosomal genotype data, for adverse pregnancy and perinatal outcomes (APPOs) via the “--glm” command using Firth logistic regression for binary outcomes and linear regression for continuous outcomes. An additive model was assumed and adjusted for the top 10 principal components of ancestry (PCs). PCs were generated using Plink –pca option after removing related individuals based on a set of independent SNPs excluding any long-range LD regions (variant count window size = 100, variant count to shift the window = 5, variance inflation factor (VIF) threshold = 1.01).

#### Born in Bradford (BiB)

***1. Overview***

Born in Bradford (BiB) is a prospective birth cohort that recruited women with expected delivery between March 2007 and December 2010 ^4^. Most women were recruited at their oral glucose tolerance test (OGTT) at approximately 26–28 weeks’ gestation, which was offered to all women booked for delivery at Bradford Royal Infirmary, except those with known diabetes, during the recruitment period. In BiB, most of the obstetric population consists of women of White British or Pakistani origin (together accounting for 81%, with the remaining women being of other ancestries). A total of 12,453 women (13,776 pregnancies) were enrolled during pregnancy who gave birth to 13,858 live children. Women had anthropometrics measured at recruitment and responded to questionnaires during pregnancy and postpartum to gain information about breastfeeding > 6 months. Biological samples were collected during pregnancy (blood and urine) and birth (cord blood) and women consented to routine primary and secondary care data linkage. Full details of the study methodology were reported previously^4^. Genetic data is available for mothers and children.

BiB contributed to this study with data on maternal genotype and perinatal outcomes (i.e., birth weight, Apgar at 1 and 5 minutes, miscarriage, stillbirth, GDM, induction of labour, caesarean section, LBW, HBW, very PTB, spontaneous PTB, post-term birth, SGA, LGA, and low Apgar at 1 and 5 minutes) that were used in the main analyses, as well as blood pressure traits during pregnancy (i.e., GH, preeclampsia, and HDP) that were used in sensitivity analyses.

***2. Genetic data***

Born in Bradford (BiB) mothers and offspring were genotyped at Bristol Bioresource Laboratories, Bristol, UK using four different Illumina arrays: HumanCoreExome12v1.0, HumanCoreExome12v1.1, HumanCoreExome24v1.0 and Infinium Global Screening Array-24 v1.0 (GSA). Genotypes were called with Illumina GenomeStudio. Genotype imputation was performed for 7606 WE and 8692 SA participants, with each ancestry and array (CoreExome/GSA) imputed separately. Prior to imputation, variants with call rate <95%, HWE P-value <1e-6 or MAF <1% were removed for each of the four ancestry/array groups separately. A/T and C/G SNPs were dropped, as were those that failed checks for strand, reference/alternative alleles, position, and SNP duplication, indels, non-autosomal SNPs, SNPs with no match in the HRC panel (position or SNP ID), and SNPs with non-matching alleles. Genotype data were imputed to the HRC r1.1 reference panel. Reference genome build version 37 (GRCh37/hg19) was used for genomic positions for BiB genotype data.

***3. Perinatal outcomes***

BiB contributed with data for the perinatal outcomes described in **Supplementary table 4**.

***4. Summary data for genotype-phenotype associations***

Genome-wide association studies (GWAS) were conducted in BiB mothers and offspring using REGENIE v3.6 to account for the complex population structure, including population stratification and relatedness, through a whole-genome regression approach. REGENIE operates in two steps: Step 1 uses array genotypes to generate 22 leave-one-chromosome-out (LOCO) predictions for each outcome, which are then included as covariates in Step 2, where imputed genotype associations are estimated. GWAS was performed separately for the two largest ancestry groups in BiB (SA and WE). The automatic phenotype imputation feature was not used for missing APPOs; instead, analyses were conducted separately for each outcome. For Step 1, called genotype data from merged arrays were used, applying PLINK 2.0 filters (“--maf 0.01”, “--hwe 0.000001”, “--geno 0.03”), resulting in 99,131 and 135,801 variants for the SA and WE samples, respectively, with a genotype block size of 1000. In Step 2, association analyses were performed using imputed autosomal genotype data, after merging samples genotyped on CoreExome and GSA arrays and retaining variants with imputation accuracy R² ≥0.3 in both arrays. A genotype block size of 200 was set, with a minor allele count (MAC) filter of 20. Linear regression was used for continuous outcomes, and Firth logistic regression (“--firth --approx --pThresh 0.999999”) was applied for binary outcomes. Standard errors were computed using the Firth regression effect size and likelihood ratio test P-value via the “--firth-se” option. Models were adjusted for genotyping array and 40 principal components: 20 internal PCs (calculated using BiB data via flashpca v2.0) and 20 external PCs (projecting BiB participants onto a UK Biobank PC space using the pcapred R package).

#### The Norwegian Mother Father and Child Cohort Study (MoBa)

1. ***Overview***

The Norwegian Mother, Father and Child Cohort Study (MoBa) is a prospective birth cohort that recruited pregnant women from all over Norway from 1999-2008 ^5 6^. The cohort includes approximately 95,200 mothers, 75,200 fathers, and 114,500 children. Mothers and their partners responded to questionnaires during pregnancy (15-, 22-, and 30-weeks’ gestation) and postpartum (6 months). Blood samples were obtained from both parents during pregnancy and from mothers and children (cord blood) at birth. Maternal anthropometrics were collected from a questionnaire at 15 weeks gestation. Data was linked to the Medical Birth Registry (MBRN), which is a national health registry containing information about all births in Norway. The current study is based on version 12 of the quality-assured data files released for research in 2019. Genetic data is available for mothers, children and partners.

2. ***Genetic data***

The generation and QC of the genetic data in MoBa have been described in full previously ^5 7^. In brief, genotyping was performed in 24 different batches on different Illumina platforms at the Genomics Core Facility, Trondheim, Norway, Erasmus MC, Rotterdam, Netherlands, and deCODE genetics, Reykjavik, Iceland. Batches that were genotyped using the same array were merged (keeping only SNPs present in all batches) and pre-imputation QC was performed on the merged batches. Principal component analysis (PCA) was used to identify participants which were genetically similar to the 1000 Genomes Project phase 1 European, Asian and African core subpopulations, after removing SNPs with MAF < 1%, call rate < 95%, and HWE p-value < 0.001. Pre-imputation QC was then performed separately for each subpopulation on the SNP, individual and family level, using PLINK 1.9 and KING 2.2.5 as the primary software packages. SNPs were removed if MAF < 0.5%, call rate < 95%, HWE p-value < 0.000001, discordant in duplicate pairs, or if associated with genotype plate and genotype batch at p-value 0.001. Individuals were removed due to being heterozygosity outliers Fhet ± 0.2 or having erroneous sex assignment, known relatedness, cryptic relatedness, identity-by-decent (PI_HAT threshold of 0.15), or being PC outliers both with and without 1000 Genomes. For family-level QC, families with more than 5% Mendel errors and SNPs with more than 1% of Mendel errors were removed, while other minor Mendel errors were zeroed out. Phasing and genotype imputation were performed using the publicly available Haplotype Reference Consortium data, via the SHAPEIT2 (duoHMM algorithm) and IMPUTE 4 software packages. Dosage data was converted to best-guess (hard call) genotype data using certainty threshold 0.7, following which low quality variants (imputation quality score [INFO] <0.8) were dropped. Post imputation QC was performed similarly to the pre-imputation QC steps, with the following changes: i) MAF filter 1%, ii) call rate <95% for SNPs, and iii) P-value for batch effects filter <5e-8. The imputation batches were then merged, and the post-imputation QC steps were repeated, using a filter for imputation batch effects rather than genotype batch effects.

3. ***Blood pressure traits during pregnancy***

Data were obtained from the Medical Birth Registry of Norway (MBRN). GH was defined as SBP≥140 mmHg and/or DBP ≥90 mmHg on at least two occasions after 20 weeks of gestation, in women who did not report having hypertension before pregnancy. Preeclampsia was defined as GH in the presence of proteinuria (≥0.3 g/d). HDP comprised both GH and preeclampsia.

4. ***Perinatal outcomes***

MoBa contributed with data for 21 perinatal outcomes described in **Supplementary table 4**.

5. ***Summary data for genotype-phenotype associations***

Summary data for genotype-phenotype associations were generated using linear/logistic regression, were conducted in MoBa mothers, fathers, and offspring using whole genome regression in REGENIE v3.1.2. Step 1 included 455,827 imputed genetic variants that were directly genotyped in at least one imputation batch and had an INFO score >0.99785. A genotype block size of 1000 was set. Step 2 involved association analyses with 6,981,748 imputed (hard-called) autosomal variants, adjusting for genotyping batch and 20 principal components. A genotype block size of 200 was used, with linear regression for continuous outcomes and Firth logistic regression (“--firth --approx --pThresh 0.999999”) for binary outcomes. The standard error was computed using the Firth regression effect size and likelihood ratio test P-value via the “--firth-se” option.

#### FinnGen

1. ***Overview***

FinnGen is the nationwide network of Finnish biobanks, which are linked to national electronic registries that provide information on prescriptions and diseases (ICD9-10 codes) ^8 9^. FinnGen includes data from 500,348 individuals (282,064 females and 218,284 males) [12^th^ data release (R12)]. Besides clinical endpoints, which also include data on some APPOs, genetic data is also available. The Coordinating Ethics Committee of the Helsinki and Uusimaa Hospital District has approved the FinnGen consortium (Nr HUS/990/2017). More information about FinnGen can be found on the website <https://www.finngen.fi/en>. The metadata from FinnGen used by the MR-PREG collaboration is publicly available at <https://www.finngen.fi/en/access_results>.

2. ***Genetic data***

Maternal genetic data were generated using Illumina and Affymetrix arrays. Genotypes were imputed using the Finnish SISu v3 reference panel. Quality control excluded samples with ambiguous gender, genotype missingness >5%, heterozygosity +-4 s.d. and non-Finnish ancestry. It also excluded variants with missingness >2%, HWE *p* <1x10-6 and minor allele count <3. Imputation was conducted using a Finnish population specific whole genome sequence reference panel.

3. ***Perinatal outcomes***

FinnGen contributed with data for all perinatal outcomes as described in **Table 2**.

4. ***Summary data for genotype-phenotype associations***

GWAS analyses were caried out in FinnGen mothers only, using whole -genome regression implemented in REGENIE version 2.2.4 (15). For REGENIE step 1, 215,152 pruned (1.5Mb window and r2 threshold of 0.2) variants were used after applying the following filters: i) imputation INFO score >0.95 in all batches, ii) >97 % non-missing genotype, and iii) MAF >1 %, and a genotype block size of 1000 was used. For REGENIE step 2, association analyses with imputed genotype data for APPOs were conducted for each variant with a minimum allele count of 5 among each phenotype’s cases and controls. The approximate Firth test was used for variants with an initial P-value of less than 0.01 and the standard error was computed based on the Firth regression effect size and likelihood ratio test P-value (REGENIE options --firth --approx --pThresh 0.01 --firth-se).

#### UK Biobank

1. ***Overview***

UKB is an adult cohort that retrospectively collected relevant data on APPOs. All people in the UK National Health Service (NHS) registry aged between 40-69 years and living within approximately 25-mile radius from one of the 22 study centres were invited to participate in UK Biobank (UKB) between 2006-2010^10 11^. A total of 500,000 adults (5.5% of the ~9.2 million invited) were recruited into the study (54.4% females). Information was assessed at baseline via a self-completed questionnaire, physical measures (including anthropometrics), and collection of non-fasting blood, urine, and saliva. Participants have been followed-up by linkage to electronic health records and a subset of participants responded to online questionnaires. Hospital Episode Statistics (HES) from 1997 for England, 1998 for Wales and 1981 for Scotland are available^12^. HES data also contain maternity-related admissions for England and Wales. It is important to note that not every participant has a hospital inpatient record, as not all have been admitted to hospital within the period covered. Genetic data from UK participants is available.

2. ***Genetic data***

Maternal genetic data were generated using the UK Biobank Lung Exome Variant Evaluation (UK BiLEVE) array (N=49,979) and the UKB axiom array (N=438,398). Imputation was performed using IMPUTE2 algorithms and a combined UK10K-HRC set as the reference panel. Quality control excluded individuals based on relatedness, gender mismatches and non-white British ancestry^13^.

3. ***Blood pressure traits during pregnancy***

Data were obtained from the Hospital Episode Statistics (HES) database. HDP were defined as SBP ≥140 mmHg and/or DBP ≥90 mmHg, measured on ≥2 occasions after 20 weeks of gestation in women who did not report having hypertension before pregnancy. This definition corresponds to codes O13 (i.e., GH) and O14 (i.e., preeclampsia) in the Tenth Revision of the International Classification of Diseases (ICD-10).

4. ***Perinatal outcomes***

UKB contributed with data for 16 perinatal outcomes, namely birth weight, gestational age, LBW, HBW, maternal depression, preterm birth, very preterm birth, post-term birth, stillbirth, miscarriage, recurrent miscarriage, sporadic miscarriage, caesarean section, emergency caesarean section, elective caesarean section and gestational diabetes. These outcomes were defined as described in **Supplementary table 4**.

5. ***Potential perinatal health risk factors***

Data on maternal age at first and last births (years), maternal height (cm), education as a proxy for socioeconomic status ("college/ university", "A level", "O level or CSEs or other", "none of the above"), parity (number of live births), BMI (kg/m^2^), smoking status (“never”, “previous” or “current” smoker) and alcohol intake frequency (“daily or almost daily”, “three or four times a week”, “once or twice a week”, “one to three times a month”, “special occasions only” or “never”) were obtained from UKB women who had ever been pregnant.

6. ***Summary data for genotype-phenotype associations***

Summary data for genotype-phenotype associations were generated using linear/logistic regressions, implemented using REGENIE version v3.6. Step 1 was performed using 549,043 directly genotyped variants with MAF ≥0.01, variant call rate ≥0.9; Hardy-Weinberg equilibrium P >1.0 x 10-15 and MAC ≥100 and set genotype block size (“--bsize”) to 1000. For step 2, association analyses were run with imputed genotype data for the APPOs, adjusting for genotyping array and 40 PCs. We set genotype block size to 200 and used linear regression or Firth logistic regression (via REGENIE options “--firth --approx --pThresh 0.999999”) for continuous and binary outcomes respectively. Standard errors were based on the Firth regression effect size and likelihood ratio test P-value as per the “--firth-se” option.

#### Other GWAS consortia for some pregnancy and perinatal outcomes

At the time of writing, the MR-PREG collaboration had harmonised and quality-controlled data from GWAS meta-analyses for GDM [5,485 cases and 347,856 controls from the GENetics of Diabetes in Pregnancy Consortium (GenDIP) consortium^14^]; PE [9,515 cases and 157,719 controls from the International Pregnancy Genetics (InterPregGen) consortium ^15^]; gestational duration-related traits [18,797 PTB cases and 260,246 controls, 15,972 post-term birth cases and 115,307 controls, and 195,555 individuals with gestation age at delivery from the Early Growth Genetics (EGG) consortium^16^]; post-natal depression [17,339 cases and 53,426 controls from the Psychiatric Genomics consortium (PGC) ^17^].

### Funding

#### ALSPAC

The UK Medical Research Council and Wellcome (Grant ref: 217065/Z/19/Z) and the University of Bristol provide core support for ALSPAC. This publication is the work of the authors and Fernanda Morales-Berstein, Gemma Clayton and Maria Carolina Borges will serve as guarantors for the contents of the paper. This research was funded in whole, or in part, by the Wellcome Trust [224982/Z/22/Z]. For the purpose of Open Access, the author has applied a CC BY public copyright licence to any Author Accepted Manuscript version arising from this submission.

A comprehensive list of grants funding is available on the ALSPAC website (<http://www.bristol.ac.uk/alspac/external/documents/grant-acknowledgements.pdf>) , but this research was specifically funded by the following grants: British Heart Foundation (SP/07/008/24066), Wellcome Trust (WT092830/Z/10/ Z and WT088806) and Lifelong Health and Wellbeing (LLHW) via the MRC (G1001357). ALSPAC GWAS data was generated by Sample Logistics and Genotyping Facilities at Wellcome Sanger Institute and LabCorp (Laboratory Corporation of America) using support from 23andMe.

#### BiB

BiB receives core funding from the Wellcome Trust (WT101597MA), a joint grant from the UK Medical and Economic and Social Science Research Councils (MR/N024397/1), British Heart Foundation (CS/16/4/32482), and the National Institute of Health Research under its Applied Research Collaboration for Yorkshire and Humber and Clinical Research Network research delivery support. Further support for genome-wide and multiple ‘omics measurements in BiB is from the UK Medical Research Council (G0600705), National Institute of Health Research (NF-SI-0611-10196), US National Institute of Health (R01DK10324), and the European Research Council under the European Union’s Seventh Framework Programme (FP7/2007–2013) / ERC grant agreement no 669545.

#### MoBa

MoBa funding is under **Acknowledgements** as requested by MoBa publication guidelines.

#### UK Biobank

UK Biobank is funded primarily by the Wellcome Trust and the Medical Research Council (MRC). It is also funded by the Department of Health, British Heart Foundation, Cancer Research UK, Diabetes UK, National Institute for Health Research (NIHR), Scottish Government, Northwest Regional Development Agency, and Welsh Assembly Government.

### Acknowledgements

#### ALSPAC

We are extremely grateful to all the families who took part in this study, the midwives for their help in recruiting them, and the whole ALSPAC team, which includes interviewers, computer and laboratory technicians, clerical workers, research scientists, volunteers, managers, receptionists and nurses.

#### BiB

BiB is only possible because of the enthusiasm and commitment of the Children and Parents in BiB. We are grateful to all the participants, teachers, school staff, health professionals and researchers who have made BiB happen.

#### MoBa

This research has been conducted using MoBa data using application number 2552. MoBa is supported by the Norwegian Ministry of Health and Care services and the Ministry of Education and Research. We are grateful to all the participating families in Norway who take part in this on-going cohort study. We thank the Norwegian Institute of Public Health (NIPH) for generating high-quality genomic data. This research is part of the HARVEST collaboration, supported by the Research Council of Norway (#229624). We also thank the NORMENT Centre for providing genotype data, funded by the Research Council of Norway (#223273), South East Norway Health Authority and KG Jebsen Stiftelsen. We further thank the Center for Diabetes Research, the University of Bergen for providing genotype data and performing QC and imputation of the data funded by the ERC AdG project SELECTionPREDISPOSED, Stiftelsen Kristian Gerhard Jebsen, Trond Mohn Foundation, the Research Council of Norway, the Novo Nordisk Foundation, the University of Bergen, and the Western Norway health Authorities (Helse Vest).

#### FinnGen

The authors thank the FinnGen investigators for sharing their summary-level data.

#### UK Biobank

We would like to thank all the participants of UK Biobank for their vital contribution to the resource. This research has been conducted using the UK Biobank Resource under Application Number 23938**.**

### Ethical approval

#### ALSPAC

Ethical approval was obtained from the ALSPAC Ethics and Law Committee and the Local Research Ethics Committees. Consent for biological samples has been collected in accordance with the Human Tissue Act (2004). Informed consent for the use of data collected via questionnaires and clinics was obtained from participants following the recommendations of the ALSPAC Ethics and Law Committee at the time (details and reference numbers of all ethics approvals can be found at <http://www.bristol.ac.uk/media-library/sites/alspac/documents/governance/Research%20Ethics%20Committee%20approval%20references.pdf>).

#### BiB

Ethical approval for the study was granted by the Bradford National Health Service Research Ethics Committee (ref 06/Q1202/48), and all participants gave written informed consent. The ALL IN sub-study had ethical approval from the London School of Hygiene & Tropical Medicine ethics committee (ref: 5320) and the Bradford Research Ethics committee (ref: 08/H1302/21). Parents (usually the mother) gave informed, written consent to take part in the study.

#### MoBa

The current study is based on version 12 of the quality-assured data files released for research in 2019. The establishment of MoBa and initial data collection was based on a license from the Norwegian Data Protection Agency and approval from The Regional Committees for Medical and Health Research Ethics. The MoBa cohort is currently regulated by the Norwegian Health Registry Act. The current study was approved by The Regional Committees for Medical and Health Research Ethics of South/East Norway (ref 2018/1256).

#### UK Biobank

The UK Biobank has approval from the North West Multi-centre Research Ethics Committee (MREC) as a Research Tissue Bank (RTB) approval. This RTB approval was granted initially in 2011 (11/NW/0382) and it is renewed on a 5-yearly cycle, with the latest one successfully renewed in 2021 (21/NW/0157).
